# Genetic insights into the biological mechanisms governing human ovarian ageing

**DOI:** 10.1101/2021.01.11.20248322

**Authors:** Katherine S Ruth, Felix R Day, Jazib Hussain, Ana Martínez-Marchal, Catherine E Aiken, Ajuna Azad, Deborah J Thompson, Hironori Abe, Jane L Tarry-Adkins, Javier Martin Gonzalez, Annique Claringbould, Olivier B Bakker, Patrick Sulem, Sandra Turon, N Charlotte Onland-Moret, Emil Peter Trane Hertz, Pascal N Timshel, Vallari Shukla, Rehannah Borup, Kristina W Olsen, Mònica Ferrer-Roda, Yan Huang, Stasa Stankovic, Paul RHJ Timmers, Thomas U Ahearn, Behrooz Z Alizadeh, Elnaz Naderi, Irene L Andrulis, Alice M Arnold, Kristan J Aronson, Annelie Augustinsson, Stefania Bandinelli, Caterina M Barbieri, Robin N Beaumont, Heiko Becher, Matthias W Beckmann, Stefania Benonisdottir, Sven Bergmann, Murielle Bochud, Eric Boerwinkle, Stig E Bojesen, Manjeet K Bolla, Dorret I Boomsma, Nicholas Bowker, Jennifer A Brody, Linda Broer, Julie E Buring, Archie Campbell, Harry Campbell, Jose E Castelao, Eulalia Catamo, Stephen J Chanock, Georgia Chenevix-Trench, Marina Ciullo, Tanguy Corre, Fergus J Couch, Angela Cox, Simon S Cross, Francesco Cucca, Kamila Czene, George Davey-Smith, Eco JCN de Geus, Renée de Mutsert, Immaculata De Vivo, Ellen W Demerath, Joe Dennis, Alison M Dunning, Miriam Dwek, Mikael Eriksson, Tõnu Esko, Peter A Fasching, Jessica D Faul, Luigi Ferrucci, Nora Franceschini, Timothy M Frayling, Manuela Gago-Dominguez, Massimo Mezzavilla, Montserrat García-Closas, Christian Gieger, Graham G Giles, Harald Grallert, Daniel F Gudbjartsson, Vilmundur Gudnason, Pascal Guénel, Christopher A Haiman, Niclas Håkansson, Per Hall, Caroline Hayward, Chunyan He, Wei He, Gerardo Heiss, Miya K Høffding, John L Hopper, Jouke J Hottenga, Frank Hu, David Hunter, Mohammad A Ikram, Rebecca D Jackson, Micaella DR Joaquim, Esther M John, Peter K Joshi, David Karasik, Sharon LR Kardia, Robert Karlsson, Cari M Kitahara, Ivana Kolcic, Charles Kooperberg, Peter Kraft, Allison W Kurian, Zoltan Kutalik, Martina La Bianca, Genevieve LaChance, Claudia Langenberg, Lenore J Launer, Joop SE Laven, Deborah A Lawlor, Loic Le Marchand, Jingmei Li, Annika Lindblom, Sara Lindstrom, Tricia Lindstrom, Martha Linet, YongMei Liu, Simin Liu, Jian’an Luan, Reedik Mägi, Patrik KE Magnusson, Massimo Mangino, Arto Mannermaa, Brumat Marco, Jonathan Marten, Nicholas G Martin, Hamdi Mbarek, Barbara McKnight, Sarah E Medland, Christa Meisinger, Thomas Meitinger, Cristina Menni, Andres Metspalu, Lili Milani, Roger L Milne, Grant W Montgomery, Dennis O Mook-Kanamori, Antonella Mulas, Anna M Mulligan, Alison Murray, Mike A Nalls, Anne Newman, Raymond Noordam, Teresa Nutile, Dale R Nyholt, Andrew F Olshan, Håkan Olsson, Jodie N Painter, Alpa V Patel, Nancy L Pedersen, Natalia Perjakova, Annette Peters, Ulrike Peters, Paul DP Pharoah, Ozren Polasek, Eleonora Porcu, Bruce M Psaty, Iffat Rahman, Gad Rennert, Hedy S Rennert, Paul M Ridker, Susan M Ring, Antonietta Robino, Lynda M Rose, Frits R Rosendaal, Jacques Rossouw, Igor Rudan, Rico Rueedi, Daniela Ruggiero, Cinzia F Sala, Emmanouil Saloustros, Dale P Sandler, Serena Sanna, Elinor J Sawyer, Chloé Sarnowski, David Schlessinger, Marjanka K Schmidt, Minouk J Schoemaker, Katharina E Schraut, Christopher Scott, Saleh Shekari, Amruta Shrikhande, Albert V Smith, Blair H Smith, Jennifer A Smith, Rossella Sorice, Melissa C Southey, Tim D Spector, John J Spinelli, Meir Stampfer, Doris Stöckl, Joyce BJ van Meurs, Konstantin Strauch, Unnur Styrkarsdottir, Anthony J Swerdlow, Toshiko Tanaka, Lauren R Teras, Alexander Teumer, Unnur Þorsteinsdottir, Nicholas J Timpson, Daniela Toniolo, Michela Traglia, Melissa A Troester, Thérèse Truong, Jessica Tyrrell, André G Uitterlinden, Sheila Ulivi, Celine M Vachon, Veronique Vitart, Uwe Völker, Peter Vollenweider, Henry Völzke, Qin Wang, Nicholas J Wareham, Clarice R Weinberg, David R Weir, Amber N Wilcox, Ko Willems van Dijk, Gonneke Willemsen, James F Wilson, Bruce HR Wolffenbuttel, Alicja Wolk, Andrew R Wood, Wei Zhao, Marek Zygmunt, Biobank-based Integrative Omics Study (BIOS) Consortium, eQTLGen Consortium, kConFab Investigators, The LifeLines Cohort Study, The InterAct consortium, Lude Franke, Stephen Burgess, Patrick Deelen, Tune H Pers, Marie Louise Grøndahl, Claus Yding Andersen, Anna Pujol, Andres J Lopez-Contreras, Jeremy A Daniel, Kari Stefansson, Jenny Chang-Claude, Yvonne T van der Schouw, Kathyrn L Lunetta, Daniel I Chasman, Douglas F Easton, Jenny A Visser, Susan E Ozanne, Satoshi H Namekawa, Joanne M Murabito, Ken K Ong, Eva R Hoffmann, Anna Murray, Ignasi Roig, John RB Perry

## Abstract

Reproductive longevity is critical for fertility and impacts healthy ageing in women, yet insights into the underlying biological mechanisms and treatments to preserve it are limited. Here, we identify 290 genetic determinants of ovarian ageing, assessed using normal variation in age at natural menopause (ANM) in ∼200,000 women of European ancestry. These common alleles influence clinical extremes of ANM; women in the top 1% of genetic susceptibility have an equivalent risk of premature ovarian insufficiency to those carrying monogenic *FMR1* premutations. Identified loci implicate a broad range of DNA damage response (DDR) processes and include loss-of-function variants in key DDR genes. Integration with experimental models demonstrates that these DDR processes act across the life-course to shape the ovarian reserve and its rate of depletion. Furthermore, we demonstrate that experimental manipulation of DDR pathways highlighted by human genetics increase fertility and extend reproductive life in mice. Causal inference analyses using the identified genetic variants indicates that extending reproductive life in women improves bone health and reduces risk of type 2 diabetes, but increases risks of hormone-sensitive cancers. These findings provide insight into the mechanisms governing ovarian ageing, when they act across the life-course, and how they might be targeted by therapeutic approaches to extend fertility and prevent disease.

## Introduction

Over the last 150 years life expectancy has increased from 45 years to 85 years^1^, but the timing of reproductive senescence (age at natural menopause) has remained relatively constant around age 50-52 years^2^.The genetic integrity of oocytes decreases with advancing age^3^ and natural fertility ceases about 10 years before menopause^4^. More women are choosing to delay childbearing to older ages, resulting in increased use of fertility treatments such as in vitro fertilisation (IVF) as well as cryopreservation of ovarian tissue or oocytes to prolong reproductive longevity^5,6^. Fertility preservation is also considered by women and girls undergoing chemotherapy and irradiation for cancer and blood disorders. However, oocyte and ovarian tissue preservation is invasive and there is a ∼6.5% chance of achieving a pregnancy with each mature oocyte thawed, but this decreases dramatically in women of advanced maternal age^7^. New methods for preserving fertility would be particularly welcome in older women.

ANM is determined by the non-renewable ovarian reserve, which is established during fetal development and continuously depleted until reproductive senescence (**Figure 1**). Follicles, consisting of oocytes and surrounding granulosa cells are formed *in utero* and maintained as resting primordial follicles in the cortex constituting the ovarian reserve. Follicles are sequentially recruited from the ovarian reserve at a rate of several hundred per month in childhood, peaking at around 900 per month at ∼15 years of age^8^. Following recruitment, follicles grow by mitotic division of granulosa cells and expansion of oocyte volume for almost 6 months until meiosis is reinitiated at ovulation and the mature oocyte is released into the oviduct. Waves of atresia (follicle death) accompany developmental transitions and growing follicles are continuously induced to undergo cell death such that, typically, only a single follicle matures to ovulate each month. As ovarian reserve declines the rate of follicle recruitment decreases, but the preovulatory follicles continue to produce substantial amounts of oestrogen, while other important hormones such as AMH and inhibin-B decline, leading to upregulation of the hypothalamus-pituitary gonadal axis.

**Figure 1.**
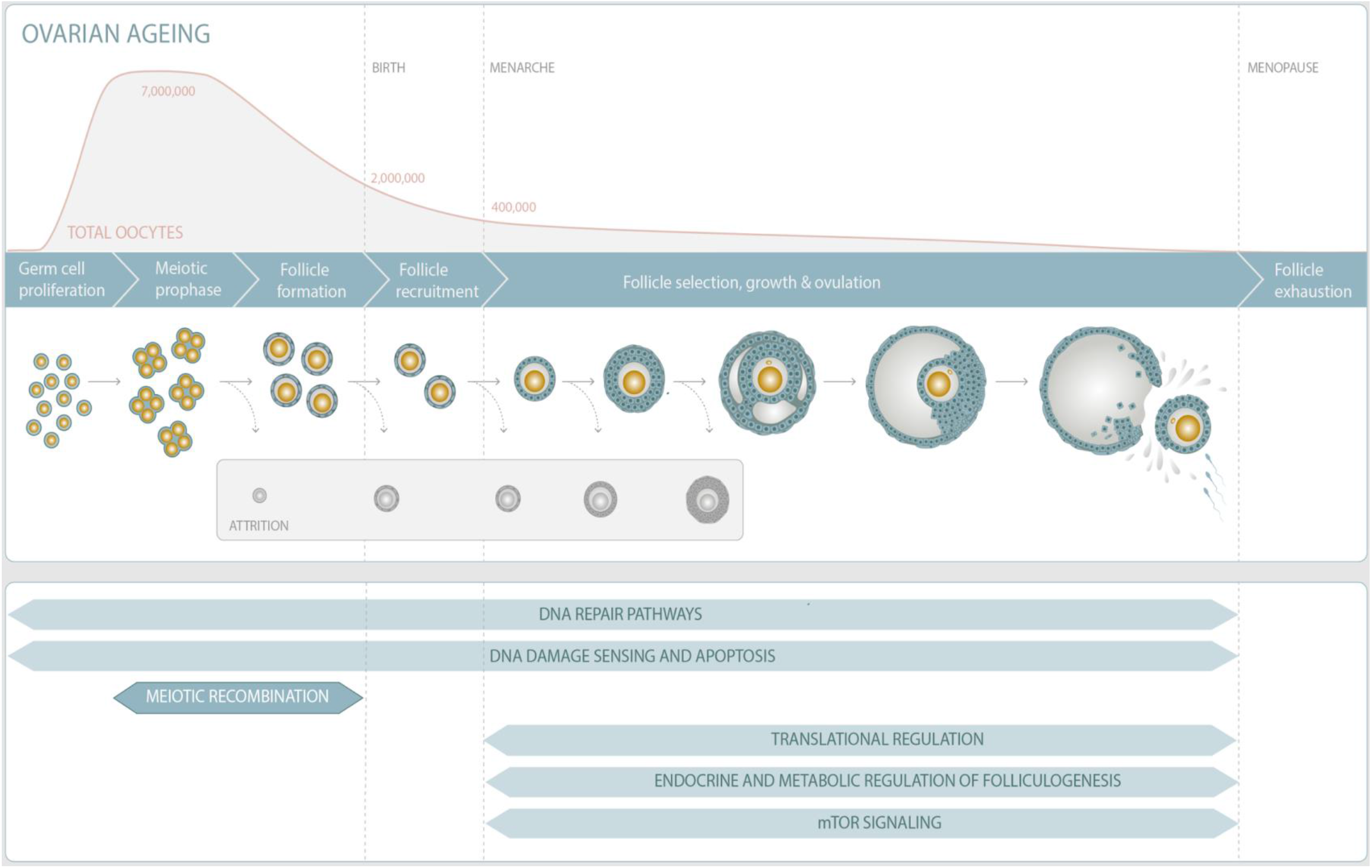
Overview of ovarian reserve and follicular activity across reproductive life.

Reproductive senescence before age 40 years is termed Premature Ovarian Insufficiency (POI) and occurs in 1 in 100 women^9^. Few lifestyle and environmental factors have been identified that contribute to variation in reproductive ageing; smoking decreases ANM^10^ and number of pregnancies is associated with increased ANM^11^. DNA damage response (DDR) is the primary biological pathway that regulates reproductive senescence, highlighted initially by genome-wide association studies (GWAS) of ANM^12^ and subsequently by rare single gene disorders that cause POI^13^ and animal models^14^. We hypothesise that DDR mechanisms could act across the life course through different mechanisms; from mitotic effects determining the peak number of primordial germ cells *in utero*, through to factors influencing the rate of follicle recruitment and cellular senescence in later life. It is not known which cell types and developmental stages are affected by genetic and environmental variation.

A better understanding of how and when molecular processes influence the establishment and decline of ovarian reserve will inform future therapeutic strategies for infertility treatment and fertility preservation. To address this, our current study describes an increase in the number of ANM-associated loci six-fold from 56 to 290, in a GWAS of ∼200,000 women, a threefold increase compared to the previous study^15^. Through subsequent integration with experiments in model organisms, we use these data to characterise the specific DDR processes that contribute to reproductive ageing, providing insights into when they act across the life-course, how they might be modified to preserve fertility and the potential consequences for broader health outcomes.

## Results

Genome-wide array data, imputed to ∼13.1 million genetic variants with minor allele frequency ≥0.1%, were available in up to 201,323 women of European ancestry (**Extended Data Fig. 1, Supplementary Table 1**). In total 38,707 genetic variants were associated with age at natural menopause (ANM) at genome-wide significance (P<5×10^−8^), which we resolved to 290 statistically independent signals (**Supplementary Table 2**). Effect estimates for these signals were highly consistent between linear and Cox proportional hazard models and across separate strata of the meta-analysis (**Extended Data Fig. 2**). This included six signals on the X-chromosome, previously untested in large-scale studies. The combined meta-analysis demonstrated no evidence of test statistic inflation due to population structure (LD score intercept = 1.02, s.e 0.03). All of the 54 previously reported^15^ HapMap 2 defined regions and the 2 exome-array signals retained genome-wide significance in this expanded dataset.

Additive, per-allele effect sizes for the 290 signals, with allele frequencies ≥0.3%, ranged from ∼3.5 weeks to ∼74 weeks (**Extended Data Fig. 1, Supplementary Table 2**). We tested all the identified signals for departure from an additive model, identifying three variants exhibiting non-additive effects (**Supplementary Table 3 and Extended Data Fig. 3**). For a common variant in *PIWIL1* (rs28416520, MAF=46%, P=2×10^−14^) a recessive model was the best fit (**Extended Data Fig. 3**). A low-frequency missense variant in *HELB* (rs75770066, MAF=3%, P=7×10^−16^) appeared to exhibit a heterozygous advantage effect (**Extended Data Fig. 3**), with higher mean ANM in the heterozygous group (95% CI 51.37-51.58 years) than the common (50.24-50.30) and rare homozygote (48.58-50.16) groups. Further fine-mapping and experimental work will be required to understand the complex biological mechanism(s) at this locus.

We next sought to validate our 290 identified signals using independent samples from the deCODE study (N=16,556 women). We observed strong directional consistency of effect estimates across individual variants (245/281, P_binomial_=2.2×10^−39^, **Supplementary Table 2**) and cumulatively these signals explained 10.1% of the variance in ANM. This compared to an estimate of 12.3% observed in UK Biobank (UKBB) samples using weights for the 290 variants derived from our non-UKBB samples (**Supplementary Table 2**). The identified signals therefore account for 31-38% of the overall genotype-array estimated heritability in UK Biobank (h^2^_g_=32.4%, s.e 0.8%), compared to 15.7-19.8% for the 56 previously reported signals. The heritability of ANM was not uniformly distributed across chromosomes in proportion to their size (**Extended Data Fig. 4**). The X-chromosome did not explain more heritability than expected given its size, however chromosome 19 explained 2.36% [1.98-2.75] of the trait variance – greater than the individual contributions of nearly all larger chromosomes (weighted average for chromosomes 1-18: 1.7%, s.e 0.2%) and ∼2.5x more than expected given its size. This was partially attributable to a single locus at 19q13 which explained ∼0.75% trait variance and where we mapped 6 independent signals (**Supplementary Table 2**).

### Common genetic variants act on clinical extremes of menopause

Elucidation of a large fraction of the underlying heritable component of ANM enables several important epidemiological questions to be addressed:

Firstly, it is unclear where in the full population distribution of ANM the influence of common genetic variants begins and ends. Our GWAS was restricted to women with ANM occurring between the ages of 40-60 years inclusive, representing ∼99% of women. ANM before 40 years (POI) is largely viewed as a Mendelian disorder, but may have a polygenic component. To test which parts of this phenotypic distribution have a polygenic basis, we calculated a polygenic score (PGS) for ANM in 108,840 women in UK Biobank using genetic weights derived from the independent non-UKBB component of the meta-analysis (**Supplementary Table 2**). The average PGS in each menopausal age group was then calculated (**Figure 2 a**), which demonstrated that ANM at a lower limit of 34 years and upper limit of 61 years had a significant polygenic influence. For example, collectively women with ANM at 34 years had an average −0.5 SD (95% CI 0.26-0.69, P=1.5×10^−5^) lower mean PGS for ANM, which we would expect to be at the population mean PGS (i.e zero) if their ANM was determined solely by monogenic variants. We had limited sample size to test outside of these age ranges, however there was some evidence for a depletion of a polygenic influence at ages younger than 34 years (**Figure 2 a**). These data suggest that common genetic variants act on clinically relevant “extremes” of ANM, although it remains unclear what fraction of POI cases may be polygenic vs monogenic.

**Figure 2.**
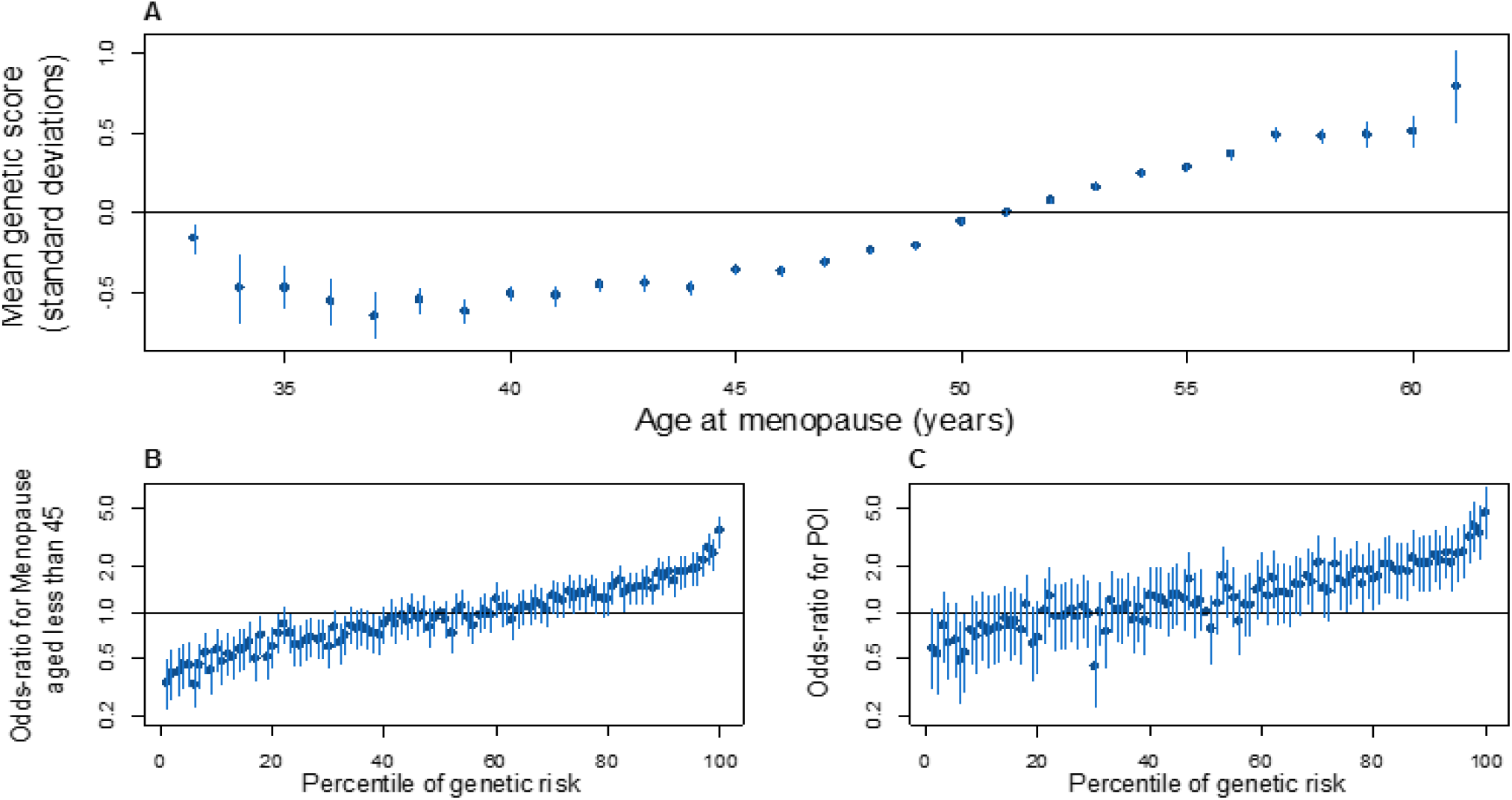
Polygenic prediction of age at menopause. (a) Changes in mean polygenic score across the spectrum of menopausal ages. Association of centile of polygenic score vs the 50th centile on b) early menopause and c) premature ovarian insufficiency.

Secondly, we evaluated the predictive ability of the ANM PGS in the independent UKBB study. Genetic risk alone proved to be a weak predictor (ROC-AUC 0.65 and 0.64 for early menopause (age <45 years) and POI respectively) (**Figure 2 b** and **c**), however the PGS performed significantly better than smoking status which is the most robust epidemiologically associated risk factor (ROC-AUC 0.58). Adding smoking status to the PGS did not appreciably improve prediction of early menopause (ROC-AUC 0.66). Despite low overall discriminative ability, the PGS was able to identify individuals at high risk of POI (**Figure 2 c**). Women at the top 1% of the PGS had equivalent POI risk (PGS OR 4.71 [3.15-7.04] vs 50th centile, P=4.4×10^−14^) to that reported for women with *FMR1* premutations, the leading tested monogenic cause of POI (OR∼5)^16^. It is however notable that the top 1% of genetic risk is more prevalent than the *FMR1* premutation carrier rate (1:250 women).

### GWAS-associated variants implicate functional genes and pathways

We used a combination of *in silico* fine-mapping and expression quantitative trait (eQTL) data integration to putatively identify the functional genes implicated by our observed genetic association signals (**Supplementary Table 2**). Firstly, 81 of the 290 independent ANM signals were highly correlated (minimum pairwise r^2^=0.8) with one or more variants that are predicted deleterious for gene function, implicating 91 unique genes. Twelve of these genes harboured predicted loss of function variants and seven genes (*MCM8, EXO1, HELB, C1orf112, C19orf57 FANCM* and *FANCA*) contained multiple statistically independent predicted-deleterious variants (**Supplementary Table 4**). We performed additional gene burden analyses for two genes with loss of function variants (*BRCA2* and *CHEK2)* using exome sequence data from 9,619 women in the UKBB study (**Supplementary Table 5**). In aggregate, women carrying loss of function variants in *BRCA2* (N=25) and *CHEK2* (N=56) reported ANM 1.69 years earlier (95% CI 0.12-3.26, P=0.03) and 2.36 years later (1.31-3.41, P=1×10^−5^) respectively (**Supplementary Table 6**). Homozygous loss of function variants in *BRCA2* were recently described as a rare cause of POI^17^, but we did not identify any such homozygotes for either *BRCA2* or *CHEK2*. Notably, the lead ANM variants mapped to within 300kb of 20/74 genes that when disrupted cause primary amenorrhea and/or POI (**Supplementary Table 7**), highlighting the common biological processes shared between normal variation in reproductive ageing and clinical extremes.

Next, we systematically integrated publicly available gene expression QTL data across 44 tissue types with our GWAS meta-analysis results (**Supplementary Table 8**). This highlighted one or more expression linked genes at 116/290 loci, which we aggregated alongside the variant mapping analyses to indicate putative functional genes at each locus (**Supplementary Tables 2 and 8**). Using three computational approaches we more broadly assessed the relative enrichment of genes expressed in each tested tissue type (**Supplementary Tables 9–14**). Hematopoietic stem cells and their progenitors showed particularly strong enrichment (**Extended Data Fig. 5**). We performed biological pathway enrichment analyses using a range of approaches, which in keeping with previous studies highlighted the importance of DDR processes as the key regulator of reproductive ageing in women (**Supplementary Tables 15–18**). We hypothesise that the shared expression profile in both haematopoietic stem cells and oocytes reflects the relative importance of DDR in both cell types^18^. In contrast to puberty timing, which represents the beginning of reproductive life, we observed no enrichment of hypothalamic and pituitary expressed genes^19^, but positive enrichment of genes expressed in the ovary and other reproductive tissues (**Supplementary Table 11**).

Finally, we attempted to leverage data from multi-tissue co-expression networks to identify genes which sit in the centre of these networks and interact with many other genes near ANM-associated variants. Such genes are analogous to the “core” genes proposed in the omnigenic model of genetic architecture^20^. This approach identified 250 genes, 47 of which were within 300kb of one of the identified 290 loci (**Supplementary Tables 19 and 20**). A notable example is *MCM8*, implicated directly by multiple predicted deleterious variants and co-expressed with many other genes highlighted by our ANM GWAS (**Extended Data Fig. 6**).

### Menopause associated genes act across the life-course

Previous large-scale genetic analyses highlighted a clear involvement of homologous recombination and the *BRCA1-A* complex in the regulation of ovarian ageing. Our current study supports much broader DDR involvement, providing increased resolution of these pathways and informing when in the life-course they might act.

Our identified genes and pathway analyses strongly implicate repair pathways associated with replication stress, in particular removal of interstrand crosslinks, which covalently join both strands of the DNA helix, as well as DNA-protein crosslinks and R loops (DNA:RNA hybrids). All of these lesions stall DNA replication and prevent transcription (**Extended Data Fig. 7)**. This observation is supported by recent work demonstrating the role of the interstrand crosslink pathway *in utero* for resolving DNA damage in pre-meiotic, primordial germ cells^21^. This process begins with replication fork remodelling at interstrand crosslinks by *FANCM*^22–24^, where we identify two independent ANM-associated missense variants (**Supplementary Table 4**). This subsequently leads to recruitment of the core Fanconi Anaemia (FA) complex to signal DNA damage, where we map missense variants in two of the eight genes – *FANCA* and *FANCB*. Furthermore we identify variants mapping key genes in the downstream repair systems coordinated by the FA pathway, including homologous recombination (e.g *RAD51, BRCA1, BRCA2*) as well as translesion synthesis (e.g *REV1, REV3L* and *RAD18*)^25^.

Several DDR genes highlighted by our study have critical meiotic functions in fetal oocytes where at least 500 programmed double-strand breaks (DSBs) initiate recombination^26^. We implicate key recombination and synaptonemal complex genes with functions in meiotic prophase (*STAG3, SMC1β, EXO1, RAD51, DMC1, HELQ, RAD52, MSH5*). Mouse models of these genes show defective repair of meiotic recombination and subsequent apoptosis of fetal oocytes resulting in decreased primordial follicles from birth and infertility^14,27–34^. We note that several of our ANM-associated variants overlap those recently reported for recombination rate^35^. This includes *NFKBIL1* and *KLHL21*, where the reported recombination rate reducing alleles were associated with later age at menopause in our data.

A range of factors likely contribute to the rate at which follicles are recruited and the follicular reserve depleted. Our data implicate key genes in the mTOR complex 1 (mTORC1) in ANM, including *STK11* and *DEPTOR*. The mTOR protein kinase that controls cell growth by regulating protein and nucleotide synthesis and is activated by the P13K pathway. Oocyte-specific deletion of *Pten* in mice removes the inhibiting effect of the PI3K pathway on primordial follicle activation, leading to premature recruitment and exhaustion of the entire primordial follicle pool^36^. Other ANM-implicated genes include *FSHB, NOBOX, INHBB, INHBC, LHCGR, IGF1, IGFBP1, PPARG* and *BMPR1B*, highlighting broader endocrine and metabolic mechanisms governing ANM. We also identified common variants in *FTO* associated with ANM which are distinct from the well-established body weight association in this region (**Supplementary Table 2**).

Finally, the majority of known genes causing POI implicate aberrant DNA damage or the inability to repair it, with limited evidence in humans that defects in the downstream cell-death signaling pathways impact variation in reproductive ageing. In contrast, our study identifies more than 58 genes implicated in regulation of apoptosis associated with ANM (**Supplementary Table 21**), providing evidence that variation in cell death following DDR is an important mechanism. This includes components and interactors of the central, conserved DDR checkpoint kinases ATR-CHEK1 (single stranded DNA) and ATM-CHEK2 (double strand breaks), that integrate and determine repair and cellular response from a broad variety of DNA repair pathways (**Extended Data Fig. 7**).

Whilst the breadth of DDR pathways identified suggests our identified loci may exert their effect at different stages across the life-course, we sought to evaluate this by assessing patterns of germ cell gene expression across different developmental stages. The individual expression profiles of our 283 highlighted genes was assessed in human fetal primordial germ cells from 5 to 26 weeks gestation, in addition to oocyte and granulosa expression in adult follicles at different stages of growth (**Extended Data Fig. 8 and Supplementary Table 22**). Collectively these data identified distinct clusters of genes that were active at different stages of life and follicle growth. The majority of our identified genes appeared most active in fetal primordial germ cells and fetal oocytes, however distinct expression profiles were evident across all developmental stages and between oocytes and granulosa cells (**Extended Data Fig. 8**). In many cases the pattern of expression was consistent with the known biological roles of those genes, for example Fanconi anemia genes were predominantly expressed in the fetal germ cells as well as the oocytes of the growing follicles, with less pronounced expression in granulosa cells (Supplementary Table 22). In contrast, genes such as *POLG* and *TP63* were predominantly expressed during follicular stages, consistent with apoptotic inducing activity in response to DNA damage observed in growing oocytes in mouse^37–40^.

### *In utero* effects may be modified by maternal diet during pregnancy

Having identified ANM associated genes as being active in the developing fetus, we next sought to evaluate whether the maternal intrauterine environment could potentially modify their expression. We used a previously described mouse model where the ovarian reserve is decreased in offspring of mothers exposed to an obesogenic diet during pregnancy^41^. Here, expression of 35 DDR genes identified in our earlier GWAS was measured in whole ovarian tissue. Of these genes, we identified significant changes in expression levels of *Dmc1* (P_adj_<0.01) and *Brsk1* (P_adj_<0.001) as a consequence of exposure to a maternal obesogenic diet during fetal life (**Extended Data Fig. 9**). *Dmc1* is a meiosis specific DNA recombinase that assembles at the site of DSBs and is essential for meiotic recombination and gamete formation^31^. We found *Dmc1* was up-regulated in the offspring of the mice exposed to the maternal obesogenic diet during fetal life. In contrast, expression levels of *Brsk1* were decreased in ovarian tissue of the offspring of these mice, an effect which appeared to be enhanced further when the offspring were additionally exposed to an obesogenic diet from weaning (**Extended Data Fig. 9**). *Brsk1* is a serine/threonine kinase that acts as a DNA damage sensor and targets *Wee1* (which also maps near one of our loci) and *Mapt1* for phosphorylation. We observed a concomitant up-regulation of expression of *Wee1* and *Mapt1* as expression of *Brsk1* was reduced (**Extended Data Fig. 9**). *Wee1* is a nuclear serine/threonine kinase that inhibits mitosis and is specifically down-regulated late in oogenesis^42^. In humans, Wee1 is expressed highly in fetal germ cells and to a lesser extent in granulosa cells of growing follicles. The mechanisms linking maternal diet-induced altered expression of these genes to reduced ovarian reserve in the offspring remain unclear. However, our findings, in addition to the observation that low birth-weight is associated with early menopause^43^, support the hypothesis that DNA repair mechanisms that act *in utero* to influence reproductive lifespan, may be modifiable by maternal exposures.

### Manipulating cell-cycle checkpoint kinase pathways extends reproductive life in animals

Previous work identified the crucial role *CHEK2* plays in culling oocytes in mouse mutants defective in meiotic recombination or after artificial induction of double-strand breaks^39,44,45^. In young females, *Chek2* inactivation can partially rescue oocyte loss and in some mutants, fertility, with high levels of non-physiologically induced endogenous and exogenous DNA damage^40,44,46,47^. To better understand the function of the checkpoint kinase pathways in physiological reproductive ageing, we used transgenic *Chek1* and *Chek2* mice (**Extended Data Fig. 10 and 11**). Follicular atresia was reduced in *Chek2*^*-/-*^ females around reproductive senescence (13.5 months). This occurred without a concomitant increase in the ovarian reserve in young mice (1.5 months) (**Extended Data Fig. 10 b**)^44^. The aged *Chek2*^*-/-*^ females showed an increased follicular response to gonadotrophin stimulation, ovulating a higher number of MII oocytes than their littermate controls, consistent with a larger ovarian reserve at 13.5 months (**Extended Data Fig. 10 c**). This was further supported by elevated anti-mÜllerian hormone levels in the aged *Chek2*^*-/-*^ females (**Extended Data Fig 10 f**). The efficiency of fertilization and blastocyst formation was not diminished by abolishing *Chek2* (**Extended Data Fig. 10 g-i**). Naturally-mated aged *Chek2*^*-/-*^ females had comparable litter sizes to wild type, suggesting that the endogenous damage that *Chek2* responds to does not compromise the health of offspring or mothers in later reproductive life (**Extended Data Fig. 10 j-k**). Thus, depletion of the ovarian reserve is slowed in *Chek2*^*-/-*^ females, resulting in improved ovarian reserve around the time of reproductive senescence and suggests a potential therapeutic target for enhancing IVF stimulation through short-term apoptotic inhibition.

In contrast to *Chek2*^−/-^, *Chek1*^−/-^ mice are embryonic lethal due to an essential function in DNA repair^48^. We found that a germline-specific conditional deletion of *Chek1* leads to defects in prospermatogonia in males^49^ and infertility in females **(Extended Data Fig. 11)**. A previous study suggests that *Chek1* is required for prophase I arrest and functions in G2/M checkpoint regulation in murine oocytes^40,50^ and that its activator, *ATR*, is important for meiotic recombination as well as follicle formation^51,52^. An extra copy, ie. three alleles, of murine *Chek1* (*SuperChek1* or *sChek1*) is reported to partially rescue lifespan in *ATR*^*Seckel*^ mice, suggesting that CHEK1 becomes rate limiting when cells are under replication stress^53^. We found that *sChek1* on its own increased the ovarian reserve from birth compared to litter-mate controls (**Extended Data Fig. 12 b, c)** as well as later in life (13.5 months; **Extended Data Fig. 12 d, e**). Large antral follicle counts were also elevated in the aged *sChek1* females, compared to litter-mate controls, indicating that follicular activity was also increased. Immediately prior to the typical age at reproductive senescence, *sChek1* females ovulated an increased number of mature MII oocytes (11-13 months, **Extended Data Fig. 12 f**), that showed similar genome stability (**Extended Data Fig. 12 g**) and had similar capacity for forming blastocyst embryos as wild type (**Extended Data Fig. 12 h, j**). When transferred, these embryos gave rise to healthy, fertile pups over two generations (**Extended Data Fig. 12 j, k, m**). Thus, *sChek1* causes a larger ovarian reserve to be established at birth and the oocytes appear to maintain their genomic integrity, as confirmed by aneuploidy analysis and efficiency of embryogenesis and fertility of pups (**Extended Data Fig. 12 g-m**), resulting in enhanced follicular activity and delayed reproductive senescence. We speculate that this is due to upregulation of replication-associated DNA repair processes during mitosis and meiosis and that repair might be limiting for establishing and maintaining the ovarian reserve. Taken together, our data show that modulating key DDR genes can extend reproductive lifespan *in vivo*, generating healthy pups that are fertile over several generations.

### Understanding the health consequences of extending reproductive life in women

After observing that DDR processes could in principle be manipulated to extend reproductive life in women, we next sought to explore what the health consequences of this might be at a population-level. To achieve this, we performed Mendelian Randomization analyses, using a number of models assessing pleiotropy, to infer causal relationships between later ANM and a range of health outcomes (**Supplementary Tables 23–25**). We focussed on outcomes that have been associated with hormone replacement therapy (HRT) in randomised controlled trials and could therefore be modifiable by extending reproductive lifespan, including metabolic traits, hormonal cancers, bone health, Alzheimer’s disease and longevity^54,55^. Consistent with previous genetic analyses and the epidemiological literature^56^, each 1-year genetically mediated later ANM increased the relative risks of several hormone-sensitive cancers by up to 5% (**Supplementary Table 23**). We note that the relationship between later ANM and higher cancer risk at the population-level is likely explained by prolonged exposure to sex hormones, while individual DDR mechansims may increase or decrease cancer risk. This conclusion is reinforced through our observation that previously reported cancer susceptibility alleles individually lead to either earlier (e.g *BRCA2*), or later (e.g *CHEK2*), ANM. Furthermore, exclusion of ANM-associated variants with larger effects on cancer risk than on ANM through Radial filtering^57^ significantly increased the strength of our MR analyses, notably for ER+ breast cancer (all ANM variants: OR=1.038, p=3.9×10^−9^, filtered: OR=1.041, p=2.7×10^−16^) (**Supplementary Table 23**). The association with prostate cancer represents a negative-control example of this concept, becoming non-significant after outlier removal (OR=1.01, p=5.5×10^−3^, changing to OR=1.00, p=0.4 after removal).

In contrast to increased cancer risks from later ANM, we observed beneficial effects of genetically-mediated later ANM on bone mineral density, fracture risk and type 2 diabetes (**Supplementary Table 23**). Our findings are consistent with evidence from randomised controlled trials that oestrogen therapy maintains bone health and protects from type 2 diabetes^55,58^. Furthermore, recent Mendelian randomisation analyses demonstrate a causal link between sex hormone levels and risk of type 2 diabetes^59^. Trial data in younger women taking HRT (50-59 years) suggested no increased risk of cardiovascular disease, stroke or all-cause mortality^55^. In agreement with this we found no evidence to support causal associations for ANM with cardiovascular disease, lipid levels, Alzheimer’s disease, body mass or longevity (**Supplementary Table 23**), all of which have been reported from non-genetic observational studies^60–66^. Finally, we evaluated putative modifiable determinants of ANM suggested by previously reported observational studies. We found that genetically instrumented increased alcohol consumption and tobacco smoking were both associated with earlier ANM. Each additional cigarette smoked per day decreased ANM by −0.05 (−0.08, −0.03) years, whilst women who drank alcohol at the maximum recommended daily limit experienced ∼1 year (−1.72,-0.26) earlier menopause compared to those who drank little. Furthermore, genetically instrumented earlier age at menarche was associated with earlier ANM (−0.15 [-0.22--0.08] years per year of earlier menarche timing) (**Supplementary Tables 26 and 27**).

## Conclusion

Collectively our analyses have provided novel insights into the biological processes underpinning reproductive ageing in women, how they can be manipulated to extend reproductive life, and what the consequence of this might be at a population level. Our identified genetic variants enabled polygenic prediction of POI which outperformed all other known existing risk factors screened clinically. Furthermore, it enabled novel insights into the causal effect of reproductive ageing on broader health outcomes in women, supporting a beneficial metabolic effect of extended reproductive life. Identified genes and pathways suggest that the genetic integrity of germ cells is the key factor determining reproductive senescence, influenced by processes across the DDR spectrum and life-course. This ranges from perturbation of DNA repair pathways regulating the amount of unrepaired DNA damage introduced during mitotic PGC expansion and meiotic recombination *in utero*, through to variability in DNA damage sensing and apoptosis pathways which impact the rate of oocyte depletion throughout life. Our mouse models suggest these pathways can be manipulated to both extend reproductive life and improve fertility, providing novel insights for potential treatments to enhance gonadotropin-stimulated oocyte retrieval in IVF. We anticipate these findings will greatly inform experimental studies seeking to identify new therapies for enhancement of reproductive function and fertility preservation in women.

## Data Availability

All genome-wide summary statistic data will be available at www.reprogen.org once the paper is published.

## ONLINE METHODS

### Phenotype definition

We included women with age at natural menopause (ANM) from age 40 to 60 inclusive. ANM was derived from self-reported questionnaire data by each study (**Supplementary Table 1**) and was the age at last naturally occurring menstrual period followed by at least 12 consecutive months of amenorrhea. Exclusions were women with menopause caused by hysterectomy, bilateral ovariectomy, radiation or chemotherapy, and those using HRT before menopause. Within each of the studies, each participant provided written informed consent and the study protocol was approved by the institutional review board at the parent institution.

### Genome-wide association study meta-analysis

A genome-wide meta-analysis of autosomal and chromosome X variants in women of European ancestry was carried out on summary statistics from analyses in three strata, allowing for the identification of heterogeneity due to different methodology. The three strata were (**Extended Data Fig. 1**): (i) meta-analysis of 1000 Genomes imputed studies; (ii) meta-analysis of samples from the Breast Cancer Association Consortium (BCAC: http://bcac.ccge.medschl.cam.ac.uk); (iii) UK Biobank GWAS. The overall meta-analysis included variants present in at least two of the three strata. All meta-analyses were inverse-variance weighted without GC correction and were carried out in METAL (https://genome.sph.umich.edu/wiki/METAL_Documentation). Analysis was conducted by analysts and two geographically distinct sites independently and the resulting summary statistics were compared for consistency.

The meta-analysis of 1000 Genomes imputed studies included 40 datasets imputed to 1000 Genomes Phase I version 3 for the autosomes and 29 for chromosome X (**Supplementary Table 1, Supplementary Notes**). Each individual study applied quality control to directly genotyped variants and samples prior to imputation (suggested exclusion thresholds for variants were Hardy-Weinberg equilibrium P<1×10^−5^, call rate <95% and minor allele frequency (MAF) <1%; suggested exclusions for samples were >5% missing genotypes, population outliers, high inbreeding coefficient, heterozygosity outliers, sex mismatches and related samples). Each individual study carried out GWAS using a two-tailed additive linear regression model adjusted for genetic principal components/relationship matrix depending on the software used (**Supplementary Table 1**), without GC correction. Since all samples included were female, chromosome X was analysed as for the autosomes. Once data were submitted, each study underwent quality control centrally according to standard protocols implemented independently by two analysts. Summary statistics for each study were stored centrally. Prior to meta-analysis, genetic variants ids were converted to “chr:position” format (position in build 37) and alleles for insertion/deletion polymorphisms were coded as “I/D” to ensure consistency across studies. Meta-analysis was carried out including SNPs with imputation quality≥0.4 and MAF≥0.001. Variants in at least half of datasets for either the autosomes or for chromosome X (as appropriate) were taken forward to the overall meta-analysis, resulting in ∼10.9 million variants.

GWAS summary statistics for the BCAC data were provided as four datasets, containing breast cancer cases and controls, with each genotyped on the iCOGs and OncoArray genotyping arrays (**Supplementary Table 1**). Quality control was applied to directly genotyped variants prior to imputation and data were imputed to the HRC r1.1 (2016) reference panel. Association analysis and quality control was carried out centrally as for the 1000 Genomes imputed studies. Summary statistics from the four BCAC datasets were meta-analysed, including variants with imputation quality≥0.4 and MAF≥0.001. Variants in two or more of the four datasets were taken forward to the overall meta-analysis, resulting in ∼14.5 million variants.

UK Biobank genotyped 488,377 participants on two arrays, 49,950 on the UK BiLEVE Axiom array (807,411 markers) and 438,427 on the UK Biobank Axiom array (825,927 markers), which were then imputed using a combined 1000 Genomes Phase 3 and HRC reference panel. Details of central genotyping, quality control and imputation are described elsewhere^67^. We included 451,454 individuals identified as European in our analysis. GWAS was carried out by applying a linear mixed model in BOLT-LMM^68^ to adjust for population structure and relatedness, also adjusting for study centre and data release. Summary statistics taken forward to the overall meta-analyses were for ∼16.6 million variants with imputation quality ≥0.5 and MAF≥0.001. UK Biobank data were analysed by two analysts independently and summary statistics results were compared for consistency.

Genome-wide significance was set at P<5×10^−8^. Statistical independence was determined using a combination of two approaches. Firstly, we used distance-based clumping to select the most significantly associated SNP within a 1Mb window. Secondly, we augmented this list with secondary signals within these 1Mb windows that were identified through approximate conditional analysis implemented in GCTA^69^. We only considered secondary signals that were uncorrelated with other selected signals (r^2^<0.05) and genome-wide significant in both univariate and joint models. 10,000 ancestry matched samples from UK Biobank were used in GCTA as an LD reference panel.

### Assessing the impact of time to event models on the signals identified

We performed Cox proportional hazards regression for the 290 genome-wide significant ANM signals, allowing inclusion in our analyses of women excluded from the definition of natural menopause. We used UK Biobank imputed genotype data and performed analyses in 379,768 unrelated individuals of European descent (as described previously), of whom 185,293 were included in our Cox analyses (phenotype definition as described previously^43^. Briefly, Cox proportional hazards regression was run using stset and stcox (Breslow method for ties) in Stata v16.0 using age as the time variable, starting at birth (0 years) and ending at last age at risk of natural menopause. Natural menopause was set as the event, with individuals censored at bilateral oophorectomy and/or hysterectomy, or start of HRT use (if ongoing at time of menopause, hysterectomy or oophorectomy). We included the covariates genotyping chip and release of genotype data, recruitment centre and the first five genetic principal components, which were considered to be constant throughout the time at risk. We calculated −1 × natural log(hazard ratio) to allow comparison with effect estimates from linear regression from the full meta-analysis and meta-analysis excluding UK Biobank.

### Confirmation of identified signals and variance explained estimates

We sought to confirm our findings by testing the 290 identified loci in aggregate in an independent sample of 16,556 women from the Icelandic deCODE study. Of those women, 14,771 were chip-typed and 1,785 are imputed 1^st^ and 2^nd^ degree relatives of chip-typed individuals. We assessed the aggregate significance of the identified loci by testing how many alleles had the same direction of effect using a binomial sign test (null expectation 50%). The proportion of variance explained using replication summary statistics provided by deCODE (n=16,556). We calculated the variance explained by each variant in deCODE (using the formula 2×β^2^×MAF×(1-MAF)), dividing the sum of the variance explained in total for the 290 variants by the SE^2^ of menopause age in deCODE

We additionally estimated the proportion of variance in ANM explained by the 290 genome-wide significant signals in UK Biobank by calculating linear regression R^2^ in 88,829 unrelated women of European descent (as described previously^70^ who had menopause age recorded. We generated estimates by combining the 290 variants as a genetic risk score with the allelic dosage weighted by the effect size from meta-analysis of the 1KG and BCAC strata only (**Supplementary Table 2**). Genotypes were extracted from imputed data and we included the covariates genotyping chip and release of genotype data, recruitment centre, age and the first five genetic principal components. Genotype-array heritability estimates were calculated using REML implemented in BOLT-LMM to provide a denominator for proportion of heritability explained.

### Assessing deviation from an additive genetic model

A dominance deviation test^71^ was run for the 290 genome-wide significant ANM signals. Briefly, in this test a dominance deviation term representing the heterozygous group (coded 0, 1 and 0) is fitted jointly with an additive genotype term in the regression model. This test determines whether the average trait value carried by the heterozygous group lies halfway between the two homozygote groups as expected under an additive model. We used best guess genotypes converted from UK Biobank imputed genotype data and performed linear regression analysis in Stata v16.0 in 379,768 unrelated individuals of European descent (identified as described previously^70^. We regressed ANM on genotype including the covariates genotyping chip and release of genotype data, recruitment centre and the first five genetic principal components. We also tested a dominant model, comparing the effect allele heterozygotes/homozygote group with other allele homozygotes, and a recessive model, comparing effect allele homozygotes with heterozygotes and other allele homozygotes. Genetic variants with a P-value for the dominance deviation term that was smaller than Bonferonni corrected P=0.05 (P=0.05/290=0.000172) were considered to show evidence of non-additive effects.

### Loss-of-function burden analyses in exome sequencing data

We analysed whole exome sequencing data from UK Biobank. Data for ∼50,000 individuals were generated externally by Regeneron and returned to the UK Biobank. We annotated variants using LOFTEE (https://github.com/konradjk/loftee) to identify individuals with high confidence loss-of-function (LoF) variants (stop-gained, splice site disrupting and frameshift variants likely to result in loss-of-function of transcript) in *BRCA2* and *CHEK2* **(Supplementary Table 5)**. Due to known issues with the exome sequencing data released by UK Biobank, we further excluded individuals/variants on the basis of allele bias (P<0.05), strand bias (P<0.05), low variant allele frequency (<0.3) in heterozygotes and low genotyping quality (<99.9%) and likely errors identified by inspecting Integrative Genomics Viewer plots of sequencing reads^72^. We calculated a burden score as the sum of LoF alleles carried by each individual in *BRCA2* and *CHEK2*. We tested the association of the burden score with ANM and incident cancers in females (all, breast, ovarian and uterine) by performing linear or logistic regression, as appropriate, in Stata v16.0. Cancer phenotypes were extracted from cancer registry data within the UK Biobank for cancers diagnosed after first study visit (ICD-9/10 codes used were: any cancer, 140–208 excluding 173/C00–C97 excluding C44; breast cancer 174/C50; ovary cancer 183/C56 and C570–C574; uterine cancer 179 and 182/C54–C55). Analyses were restricted to samples identified as being unrelated and of European descent (identified as described previously^70^). Recruitment centre, age (for cancers only) and the first five genetic principal components were included as covariates. In the same way, we repeated this analysis for the *CHEK2* 1100delC loss-of-function variant (rs555607708, GRCh38 chr22:28695868:AG>A) since this variant is included in the LoF burden score but is more common than other such variants (MAF 0.2%).

### Identifying putatively functional genes

We used two in silico approaches to prioritise putatively functional genes across our highlighted loci. Firstly, To identify variants with functional consequences, we looked up variants in r^2^>0.8 with the signals in Variant Effect Predictor (build 38). We identified missense, frameshift, insertion/deletions and stop-gained and splice site disrupting variants, which we then classified according to their VEP, PolyPhen and SIFT impact. We considered ‘high impact’ variants as those classified as high impact by VEP (stop-gained, frameshift and splice site disrupting). ‘Medium impact’ variants were missense variants classed as moderate impact by VEP, which were either deleterious in SIFT and were at least possibly damaging in PolyPhen. ‘Low impact’ variants were missense or inframe insertions/deletions classed as moderate impact by VEP and were tolerated and/or benign in PolyPhen. LD was calculated using PLINK v1.9 from best guess genotypes for 1000 Genomes Phase 3/HRC imputed variants in ∼340,000 unrelated UK Biobank participants of white British ancestry. Genetic variant locations were converted from b37 to b38 using UCSC Liftover.

Secondly, we integrated our ANM genome-wide summary statistics with eQTL data using Summary Mendelian Randomization (SMR)^73^. Publicly available expression datasets for 48 tissues in GTEx v7 and 10 brain regions were downloaded from the SMR website (https://cnsgenomics.com/software/smr/#eQTLsummarydata). Whole-blood data in an eQTL meta-analysis of 31,684 samples was available from the eQTLGen consortium [https://www.biorxiv.org/content/10.1101/447367v1] A Bonferroni corrected p-value threshold was used in each expression dataset individually and only associations with HEIDI P > 0.01 were considered to avoid coincidental overlap due to extended patterns of LD. This resulted in a total of 44 (SMR P<7×10^−6^) significant transcriptions in the brain, 96 in whole blood (P<3×10^−6^) and 732 across all GTEx tissues (SMR P<3.6×10-7). We excluded brain and whole blood tissues from the collection of 48 tissues in GTEx as they were better represented by the other expression datasets.

### Identifying enriched cell and tissue types

We used three approaches to identify cell and tissue types enriched for ANM associated variants. DEPICT was run using default settings as described previously^74^, using GWAS summary statistics including all autosomal variants with P-value <1×10^−5^. The cell-type specific expression matrices used as input to DEPICT were generated from individual single-cell gene expression datasets (see below). Briefly, each data set was processed by first normalizing cells’s gene expression to a common transcript count (10,000 transcript per cell) before calculating the average expression of each gene for each cell-type annotation. Averaged data was log-transformed (natural log). We computed cell-type specific gene expression following using a two-step z-score approach - first we calculated gene-wise z-scores (each gene; mean=0, sd=1) to remove the effect of ubiquitous expressed genes, then we calculated cell-type-wise z-scores (each cell-type; mean=0, sd=1) on gene-wise z-scores. For mouse expression datasets we mapped mouse genes to human orthologs using Ensembl (v. 91) keeping only genes with a 1-1 ortholog mapping.

DEPICT analyses were run on four datasets: 1) Tabula Muris (https://tabula-muris.ds.czbiohub.org/)^75^, restricted to the fluorescence-activated cell sorting samples. To keep the tissue level information in the dataset, we defined cell-type annotations as ‘tissue cell-types’ by combining the cell-type label (‘cell_ontology_class’ column) with the origin tissue of the cell-type (‘tissue’ column). This allowed us to e.g. distinguish B-cells originating from fat, spleen and marrow tissue. In total we analyzed 115 cell-type annotations from 44,949 cells; 2) Zeisel et al. Mouse brain dataset^76^ covered 265 cell-type annotations derived from 160,796 cells; 3) Nestorowa et al. human hematopoietic stem and progenitor cell differentiation dataset^77^ was not normalized to a common transcript count because the data was pre-normalized by the authors. We defined cell-type annotations as the 12 distinct hematopoietic stem and progenitor cell (HSPC) phenotypes reported by the authors (shown in their manuscript Figure 3A). The annotations covered 1,483 cells; 4) the Baron et al. pancreas dataset^78^ from four human pancreatic samples covered 14 cell-type annotations from 8,569 cells.

Secondly, we additionally performed tissue enrichment analysis using linkage-disequilibrium (LD) score regression to specifically expressed genes (LDSC-SEG)^79^. We used three datasets available on the LDSC-SEG resource page (https://github.com/bulik/ldsc/wiki/Cell-type-specific-analyses), relating to cell and tissue-specific annotations from GTEx^80^, Epigenome Roadmap^81^ and the “Franke lab”^74,82^.

Finally, tissue enrichment analyses were performed using ‘Downstreamer’, which is described in a separate section below.

### Pathway analysis

MAGENTA was used to explore pathway-based associations in the full GWAS data set. MAGENTA implements a gene set enrichment analysis (GSEA)-based approach^83^. We used upstream and downstream limits of 110Kb and 40Kb to assign variants to genes, excluded the HLA region from the analysis and set the number of permutations to 10,000 for GSEA testing, with analysis using 75% and 95% cut-offs. Significance was determined when an individual pathway reached FDR<0.05 in either analysis. In total, 3,222 pathways from Gene Ontology, PANTHER, KEGG and Ingenuity were tested for enrichment of multiple modest associations with ANM.

We additionally performed pathway analyses in ‘Downstreamer’ (described in section below) and MAGMA v1.08^84^. MAGMA analyses were performed using the full genome-wide summary statistics, but restricted to variants that were predicted deleterious (i.e non-synonymous and loss of function). Gene-sets included in the analyses were obtained from MsigDB v7.2, which included 12,358 curated gene sets from KEGG, Reactome, BioCarta and GO terms consisting of biological processes, cellular components and molecular functions.

## Downstreamer methodology

### Calculation of GWAS gene p-values

Downstreamer works by using gene p-values that are derived from the GWAS variant p-values. First we applied genomic control to correct for inflation in the GWAS signal. We then integrated the procedure from the PASCAL^85^ method into Downstreamer so that we can sum variant p-values into a gene p-value while accounting for the local linkage disequilibrium (LD) structure. We used all variants within a 25kb window around the start and end of a gene and we used the non-Finnish European samples of the 1000 Genomes (1000G) project, Phase 3 to calculate LD^86^. We then calculated these GWAS p-values for all 20,327 protein-coding genes (Ensembl release v75).

### Null GWAS to account for chromosomal organization of genes and empirical p-value calculations

We used randomized null GWAS for steps in the Downstreamer methodology. To do so, we first simulated random phenotypes by drawing phenotypes sizes from a normal distribution and then associated them to the genotypes of the 1000G Phase 3 non-Finnish European samples. Here we only use the overlapping variants between the real traits and the permuted GWASes. The simulated GWAS signals are random and independent of each other, so the remaining correlation between gene p-values reflects the underlying LD patterns and chromosomal organization of genes. We simulated 10,000 GWAS in this way and used these to calculate correlations between genes under the null. The next 10,000 simulated GWASes are used to empirically determine enrichment p-values and finally we used an additional 100 simulations to estimate the false discovery rate (FDR) of Downstreamer.

### Pre-processing of GWAS gene p-values

For each GWAS, both real and simulated, we force normalized the GWAS Z-scores into a normal distribution to ensure that outliers would not have disproportionate weights. Due to limitations in the PASCAL methodology that result in ties at a minimum significance level of 1×10^−12^ for highly significant genes, we use the minimum SNP P-value from the GWAS to identify the most significant gene. We then used a linear model to correct for gene length, as longer genes will typically harbour more SNPs.

Sometimes, two (or more) genes will be so close to one another that their GWAS Z-scores are highly correlated, violating the assumptions of the linear model. Thus, genes with a Pearson correlation r ≥ 0.8 in the 10,000 GWAS permutations were collapsed into ‘meta-genes’ and treated as one gene. Meta-gene Z-scores were averaged across the input Z-scores. Lastly, the GWAS Z-scores of the meta genes were scaled (mean = 0, standard deviation = 1).

### Pathway and gene set gene prediction

To identify pathway and disease enrichments, we used the following databases: Human Phenotype Ontology (HPO), Kyoto Encyclopaedia of Genes and Genomes (KEGG), Reactome and Gene Ontology (GO) Biological Process, Cellular Component and Molecular Function. We have previously predicted how much each gene contributes to these gene sets, resulting in a Z-score per pathway or term per gene^87^. We collapsed genes into meta-genes in parallel with the GWAS step, to ensure compatibility with the GWAS Z-scores following the same procedure as in the GWAS pre-processing. Meta-gene Z-scores were calculated as the Z-score sum divided by the square root of the number of genes. Finally, all pathway Z-scores were scaled (mean = 0, standard deviation = 1).

### Co-regulation matrix

To calculate core scores, we used a previously generated co-regulation matrix that is based on a large multi-tissue gene network^87^. In short, publicly available RNA-seq samples were downloaded from the European Nucleotide Archive (https://www.ebi.ac.uk/ena). After QC, 56,435 genes and 31,499 samples covering a wide range of human cell-types and tissues remained. We performed a PCA on this dataset and selected 165 components that offered the best prediction of gene function. We then selected the protein coding genes and centred and scaled the eigenvectors for these 165 components (mean = 0, standard deviation = 1) such that each component was given equal weight. The first components mostly describe tissue differences^87^, so this normalization ensures that tissue-specific-patterns do not disproportionately drive the co-regulation matrix. The co-regulation matrix is defined as the Pearson correlation between the genes from the scaled eigenvector matrix. The diagonal of the co-regulation matrix was set to zero to avoid these genes having a disproportionate effect on the association to the gene P-values. Finally, we converted the Pearson r to Z-scores.

### Generalized least squares model to calculate pathway enrichment and core gene scores

We used a generalized least squares (GLS) regression to associate the GWAS Z-scores to the pathway Z-scores and co-regulation Z-scores. These two analyses result in the pathway enrichments and core gene prioritisations, respectively. We used the gene-gene correlation matrix derived from the 10,000 permutations as a measure of conditional covariance of the error term (**Ω**) in the GLS to account for the relationships between genes due to LD and proximity. The pseudo-inverse of **Ω** is used as a substitute for **Ω**^−1^

The formula of the GLS is as follows:

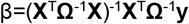

Where β is the estimated effect size of pathway, term or gene from the co-regulation matrix, **Ω** is the gene-gene correlation matrix, **X** is the design matrix of real GWAS Z-scores and **y** is the vector of gene Z-scores per pathway, term or gene from the co-regulation matrix. As we standardized the predictors, we did not include an intercept in the design matrix and **X** only contains one column with the real GWAS Z-scores. We estimated the beta’s for the 10,000 random GWASes in the same way and subsequently used them to estimate the empirical p-value for β.

## Definition of POI and DDR genes

We combined genes implicated in the DDR from a number of sources yielding a total of 778 genes (**Supplementary Table 21**)^88–90^. To identify genes associated with premature ovarian insufficiency/primary ovarian insufficiency (ICD-11 GA30.6), we carried out a search in PubMed for premature ovarian insufficiency, primary ovarian insufficiency, premature ovarian failure and ovarian dysfunction in humans and reviewed all primary studies published in English until 22nd of July, 2020. We included syndromic, non-syndromic, sporadic as well as familial single nucleotide variants, insertion/deletions and copy number variants (CNVs) and included 114 genetic variants from 139 studies. We did not attempt to review the clinical significance of the variants, which ranged from classical POI genes to newly identified CNVs in whole-exome sequencing studies. We expanded our search to review articles and ClinVar. We uncovered another four genes implicated in Perrault Syndrome for which our search terms were not included in the original articles. This gave a total of 118 genes. Our search detected all genetic variants entered in ClinVar as pathogenic, likely pathogenic or with conflicting interpretations of pathogenicity. We excluded genes with variants when no assertion criteria were provided and no published data were available for assessment in ClinVar. Two studies of large chromosomal rearrangements as well as quantitative trait loci consisting of more than a single genetic variant from GWAS in POI populations were excluded resulting in 74 genes (**Supplementary Table 7**).

## Polygenic prediction of early menopause

To evaluate the impact of common variants on clinical extremes of ANM, we first performed a GWAS meta-analysis excluding the UK Biobank study (N=95,275). Effect estimates from this analysis (**Supplementary Table 2**) were then used for subsequent polygenic risk score (PRS) construction of ∼6.97 million autosomal variants across the genome using LDPRED^91^. The PRS was calculated using PLINK^92^ v1.90b4.4 in an independent sample of 108,840 women from the UK Biobank study, rescaled to have a mean of 0 and standard deviation of 1. We then estimated the centile distribution of the genetic risk score for all women with a valid ANM (with no lower or upper phenotype boundary). Two outcomes were defined: early menopause (EM) defined as ANM < 45 (N=11,268) vs all other women (N=97,572); and premature ovarian insufficiency (POI), defined as ANM < 40 years (N=2,407) vs all other women (N=106,433). Logistic regression analyses, adjusting for age, genotype array and 10 genetic principal components, were then performed with either EM or POI as the outcome. This was performed 99 times for each centile of genetic risk (coded 1) vs the 50^th^ centile of genetic risk (coded 0). To assess the relevance of this score to each ANM age group, we estimated the average PRS value by year of ANM. For example, we grouped all women with ANM = 47 and estimated the mean and standard error of the PRS in this group of women. Our intuition was that any ANM range not influenced by common genetic variants would have the population mean PRS (i.e mean = 0 and SD = 1). Receiver operating characteristics (ROC) models were performed in Stata v14 using the *roctab, rocgold* and *rocreg* commands.

## Mendelian Randomization analyses

In order to infer causal relationships between ANM and other health related outcomes, we performed Mendelian Randomization (MR). The 290 independent ANM signals were used as a genetic instrument for later ANM. Where a signal was not present in the outcome GWAS, we identified the best HapMap2 proxy with r2>0.5 within 250 kb either side of the signal and its relevant weight was included in our genetic instrument (**Supplementary Table 25**). The genetic variants were identified in publicly available GWAS datasets for a range of outcomes of interest (**Supplementary Table 24)**. These were used in three methods of MR - inverse variance weighted^93^, MR-EGGER^94^ and weighted median^95^. As a sensitivity analysis we additionally removed signals that appeared to be outliers. This was achieved using the Radial method considering the IVW model^57^. We also performed MR considering the effect of a range of putative modifiable risk factors on ANM as the outcome using the same MR models. Genetic instruments were created for the risk factors using independent genetic variants with effects estimated in published GWAS (**Supplementary Table 27**). For the risk factors of cigarette exposure and alcohol consumption, the MR was performed with a single genetic variant by calculating a Wald ratio for the effect of the variants on ANM divided by the effect on the risk factor using mrrobust in Stata v16.0. The effect of the genetic variant for alcohol consumption was measured in log(drinks per week) (note that drink is a US measure of alcohol consumption equal to 14g pure alcohol, equivalent to 1.75 UK units). Hence a change from 1 drink to 7 drinks (US maximum recommended per week) would be the equivalent of a 1.95 increase in log(drinks per week), which when applied to the Wald estimate, gives the respective change in age at menopause.

### Expression of candidate genes identified by human GWAS in a mouse model of environmentally-induced low ovarian reserve

#### Generation of mouse model

All animal experiments underwent ethical review by the University of Cambridge Animal Welfare and Ethical Review Board and were carried out under the UK Home Office Animals (Scientific Procedures) Act (1986, United Kingdom). Female C57BL/6J mice were randomized to be fed *ad libitum* either a standard laboratory chow diet (7% simple sugars/3% fat; Special Dietary Services, Witham, UK) or an obesogenic diet (10% simple sugars/20% animal lard; Special Diets Services, Witham, UK). The obesogenic diet was supplemented with a separate pot of sweetened condensed milk (55% simple sugars/8% fat; Nestle UK, Gatwick, UK) available to the animals within the cage. A detailed description of the dietary regimen has been published previously^96^. Female mice were placed on the allocated diet six weeks prior to first mating with wild-type males on standard chow diet. The first litter was discarded after weaning, and only proven-breeder females were used for the experimental protocols. Second matings occurred once females on the obesogenic diet had reached at least 10g absolute fat mass, as assessed by time domain nuclear resonance imaging (TDNMR) (Minispec Time Domain Nuclear Resonance, Bruker Optics). The female mice remained on their allocated diets throughout the breeding, pregnancy, and lactation phases. After delivery, each litter was culled to six pups at random to standardize their plane of nutrition from postnatal day 3 in all litters. There was no significant difference in the pre-culling litter size between obesogenic and control litters. Equal sex ratios within the litters were maintained as far as possible. After weaning at day 21, female offspring were randomly allocated to either the control or the obesogenic diets (identical to those used for the dams) and remained on these diets for the duration of the study. Bodyweight and food intake were measured weekly. At 12 weeks of age, offspring total and fat mass were assessed by weighing and by TDNMR (Minispec Time Domain Nuclear Resonance, Bruker Optics) respectively. Following an overnight fast, the female offspring were weighed and then culled by CO_2_ asphyxiation and cervical dislocation. Ovaries were dissected and weighed immediately. One ovary from each animal was snap-frozen in liquid nitrogen or dry ice, and stored at −80^°^C, the other was fixed in formalin/paraldehyde. The fixed ovary was sectioned and subjected to haematoxylin and eosin (H&E) staining to ensure equal distribution of estrous stages in each experimental group (data not shown). Detailed reproductive and metabolic phenotyping of the female pups has previously been published^41^.

#### Gene expression analysis

A screen of 35 DNA damage response genes highlighted by our previous GWAS on ANM were selected for investigation^15^ - Brca1, Bre, Brsk1, Chd7, Chek2, Dido1, Fbxo18, Helb, Helq, Mcm8, Mff1ip, Msh5, Msh6, Mycbp, Polg, Prim1, Rad51, Rad54, Rev3l, Uimc1, Apex1, Aptx1, Cdk2ap1, Dmc1, Exo1, Fam175a, Fanci, Ino80, Kntc1, Papd7, Parl, Parp2, Polr2e, Polr2h and Tlk1. Expression levels were measured in whole snap-frozen ovaries. RNA was extracted using a miRNeasy mini Kit (Qiagen, Hilden, Germany). The kit was used according to the manufacturer’s instructions, with the addition of DNaseI digestion to ensure that the samples were free from genomic DNA contamination. The extracted RNA was quantified using a Nanodrop spectrophotometer (Nanodrop Technologies, Wilmington, DE, US). cDNA was synthesized from 1μg RNA using oligo-dT primers and M-MLV reverse transcriptase. Gene expression was quantified via RT-PCR (StepOne Plus machine; Applied Biosystems, Warrington, UK) using custom-designed primers (Sigma, Poole, UK) and SYBR green reagents (Applied Biosystems, Warrington, UK). Equal efficiency of reverse transcription between all groups was confirmed using the housekeeper gene *ppia*, and absence of gDNA contamination was confirmed by quantifying *myh6*, which was absent in all samples.

#### Statistical analysis

All data were initially analyzed using a 2-way ANOVA with maternal diet and offspring diet as the independent variables. In order to correct for multiple hypothesis testing of gene expression levels, p values were transformed to q values to take account of the false discovery rates using the p.adjust function in R stats package (R Foundation for Statistical Computing, Vienna, Austria). Data are represented as means ± SEM. Where p values are reported, an alpha level <0.05 was considered statistically significant. All data analysis was conducted using the R statistical software package version 2.14.1 (R Foundation for Statistical Computing, Vienna, Austria). In all cases, n refers to the number of litters, and n=8 for all groups. Study power was determined based on effect sizes for gene expression differences observed in our previous studies of this model^41^.

### Human oocytes mRNA screen

Research on RNA expression in human eggs was carried out according to the Helsinki II declaration and was conducted in accordance with national regulation on research on human subjects and material. The research was approved by the Scientific Ethical Committee of the Capital Region of Denmark (Videnskabsetisk Komite) in accordance with Danish National regulation (H-2-2011-044; extension license amm. Nr. 51307; license holder: Claus Yding Andersen and H-1604473; license holder: Eva R. Hoffmann; H-16027088 granted to Marie Louise Grøndahl). GDPR approval was obtained from the national data agency (SUND-2016-60, Eva R Hoffmann and HGH-2016_086 to Marie Louise Grøndahl). Single human MII oocytes were collected as described previously^97^, lysed in-tube and the cDNA was amplified according to the manufacturer’s instructions (Takara Bio; mRNA-Seq, SMART-Seq v4 ultra low input RNA kit, cat. no. 634894). The quality of individual cDNA libraries was verified on an Agilent 2100 Bioanalyzer instrument using a high sensitivity DNA kit (Agilent, 5067-4626). The libraries were prepared with 100 pg input using the Nextera XT DNA library preparation kit (Illumina, FC-131-1024) and the Nextera XT index kit v2 (FC-131-2002) and quantified on a Qubit 3.0 fluorimeter (Thermo Fisher Scientific, Q32854). The quality of the final library was verified on the Agilent 2100 Bioanalyzer high sensitivity DNA chip and pooled to 4 nM. The 4 nM library pools were denatured and loaded according to the recommended NextSeq500 guidelines (Illumina Inc.).

### sChek1, Chek1 cKO, and Chek2 mice

Mouse work at the University of Copenhagen (*sChek1*) was licensed under 2016-15-0202-00043 by the Danish Animal Experiments Inspectorate (Dyreforsøgstilsynet, Denmark). Mouse work at UAB (Chek2) was approved by the UAB and the Catalan Ethics Committee for Animal Experimentation (CEEAAH 1091; DAAM6395). Mouse work at CCHMC (Chek1) was performed according to the guidelines of the Institutional Animal Care and Use Committee (protocol no. IACUC2018-0040) approved by CCHMC.

*Chek1 cKO, Chek2* mutant mice were generated previously^49,53,98^. The lines were maintained in a C57BL6 (*Chek1cKO)* or C57BL/6-129Sv (*Chek2 and sChek1*) mixed background, respectively. All experiments were carried out using litter mate controls or with animals of closely related parents as controls. The three mutant strains were kept at the University of Copenhagen (*sChek1*), Autonomous University of Barcelona (*Chek2*), and Cincinnati Children’s Hospital Medical Center (*Chek1 cKO*). Breeding cages were set in a conventional way with strict specific pathogen-free barrier and mice used for experiments were kept in individual ventilated cages (IVC). 12h light exposure was provided. Temperature, relative humidity and air changes per hour were 22 °C (+/-2 °C), 55% +/-10 %, and 17 respectively. Food and water were provided *ad libitum*. Animals were genotyped two times, initially upon weaning and again before experimental procedures were carried out. Mouse genotyping was performed by PCR analysis using the following primers for *sChek1*: gsChek1_left “TGT CTT CCC TTC CCT GCT TA”, gsChek1_right1 “TCC CAA GGG TCA GAG ATC AT” and g*s*Chek1_5’PCR2 “GTA AGC CAG TAT ACA CTC CGC TA”. The wild type gene yields a size of 400 bp whereas the transgene is 270 bp. For *Chek2*, the primers WT1F (5’–GTGTGCGCCACCACTATCCTG–3’), WT2R (5’–CCCTTGGCCATGTTTCATCTG–3’) and NeoMutR (5’–TCCTCGTGCTTTACGGTATC–3’) were used to detect the wild type (450 bp) and the mutant (625 bp) alleles in one PCR reaction. The Qiagen Taq polymerase PCR kit was used for genotyping (Cat No 201203 / 201205). All animals were sacrificed using CO_2_ following the protocols approved by the ethics committees. All subsequent protocols for mouse experiments were the same across the two participating Centers.

### Mouse ovarian histology and follicle count

Ovaries were dissected and placed in Bouin’s fixative solution (70% saturated picric acid solution (Applichem, A2520, 1000), 25% formaldehyde, 5% glacial acetic acid (Merck, 1.00063.2500)) and fixed overnight at 4 °C. The ovaries were washed two times with cold PBS for 30 minutes followed by dehydration with an increasing concentration of ethanol.

Subsequently, the samples were submerged in Histo-Clear II (Cat. # HS-202, National Diagnostics) for 30 min. at room temperature. This was repeated another two times (three times in total) with fresh Histo-Clear II. Ovaries were embedded in paraffin blocks and cut to a thickness of 7 µm and mounted on poly-L-lysine coated slides. After de-paraffinization and rehydration, the slides were stained with PAS-hematoxylin. The tissue was imaged using a Zeiss Axio scanner Z.1 and follicles with a visible nucleus were counted using the Zen Blue lite software from Zeiss. Primordial follicles contain one layer of flat granulosa cells surrounding the oocytes, primary follicles have one layer of cuboid granulosa cells. Secondary follicles contain two or more layers of granulosa cells and antral follicles are those with one or several cavities (the antrum).

### Mouse ovulation induction and oocyte collection

Ovulation was induced by injection of 5 IU of PMSG (Prospec; ref HOR-272) followed by 5 IU of hCG (Chorulon Vet; ref 422741) after 47 hours. For 11-13, 16 and 24 months old mice, 7.5 IU of each hormones were used. 12 hours post-hCG injection, the mice were sacrificed and oviducts were dissected under a stereo-microscope to release the cumulus masses into 90 µl drop of fertilization medium covered with mineral oil (NordilCell; ref 90142). Oocytes were recovered from oviducts by gently tearing swollen ampulla of oviducts to release cumulus masses into medium. Recipe of fertilization medium was previously published elsewhere^99^.

### Mouse embryo development in vitro

Pre-thawed frozen sperm from a proven fertile, C57BL/6N wild-type male was used for *in vitro* fertilization and poured into a dish containing mature MII eggs in fertilization medium. Disappearance of germinal vesicle (GV) and polar body extrusion confirmed fertilization. Zygotes were incubated at 5% CO_2_ and 37 °C. After incubating zygotes in fertilisation medium for overnight, We transferred zygotes to a 60 mm petri dish containing 50 µl KSOM (Chemicon, cat MR-106-D) covered by mineral oil(NordilCell; ref 90142). Two separate dishes were prepared for embryos from each genotype. The embryos were again incubated at 5% CO_2_ and 37 °C. The developmental stage of embryos was assessed using a stereomicroscope at the equivalent of 0.5, 1.5, 2.5, 3.5, 4.4 and 5.5 days post-coitum (dpc).

### Mouse embryo transfer

Wild-type female recipient mice (surrogate) were prepared to receive embryos by mating them with an infertile male one night before the transfer of embryos. Successful preparation of recipient mice for embryo transfer was confirmed by checking for the presence of a plug. Two cell-stage (1.5dpc) embryos were transferred into a single horn of recipient mice and anaesthesia were maintained during this procedure. Pups were born after 19 days of embryo transfer.

### Natural breeding, assessment of health of offspring and fertility in mouse

To test the natural breeding efficiency, we set cages with one or two adult (2-months or 12-month-old) control or females with a male of proven fertility. We registered litter sizes and dates of delivery for all litters obtained during a period for up to one year.

### Mice Serum AMH analysis

Mice of various ages were anesthetized. Blood was collected in a plain tube, allowed to clot for one hour at room temperature and then centrifuged at 3000 rpm (1500g) for 15 minutes at 4 °C. After centrifugation, supernatant (serum) was collected in a 1.5 ml tube and stored at −80 °C. Serum AMH levels were determined by using AMH ELISA kit (cat. # AL-113) from Ansh Labs, Webster, TX.

Assessment of the health of the offspring from control and mutant breeding was performed on a weekly basis by the personnel of the respective animal facilities following the standard health monitoring protocols approved by the Copenhagen or Catalan Ethics Committee for Animal Experimentation.

### Expression analysis of GWAS genes in human oocytes and granulosa cells at various stages of development

We used processed RNA-seq data of Fetal Primordial Germ Cells from Li *et al* (2017, Accession code: GSE86146)^100^ from 17 human female embryos ranging from 5-26 weeks post-fertilisation, and from Zhang *et al* (2018, Accession code GSE107746)^101^ studies, follicles at 5 different stages of development from fresh ovarian tissue from 7 adult donors, separated into oocytes and granulosa cell fractions; in addition to our MII Oocytes single-cell RNA-seq dataset (described below).

We transformed the per-cycle base call (BCL) file output from the sequencing run of 11 human MII oocytes into per-read FASTQ files using the bcl2fastq2 Conversion Software v2.19 from Illumina. The samples libraries were multiplexed across four sequencing lanes and the FastQ files from each of the four lanes were concatenated to generate one set of paired fastq files per sample. We performed sample QC and filtering of reads to remove low quality reads, adaptor sequences and low quality bases with trimmomatic version 0.36^102^ in two steps using ILLUMINACLIP:/ /Trimmomatic-0.36/adapters/NexteraPE-PE.fa:2:30:10 (SLIDINGWINDOW:4:20 CROP:72 HEADCROP:10 MINLEN:40 followed by and extra trim of headbases with HEADCROP:10.) Subsequent to filtering, we used the remaining paired reads for alignment by hisat2^103^ to the human genome GeneCode v.27 release with the paired GenCode v.27 gtf file containing gene annotations using: ($HISAT2 -p 22 --dta -x.gencode.v27 −1 R1.fastq −2 R2.fastq -S sample.sam) (Pertea *et al*. 2016). The resulting sam files were sorted, indexed and transformed to bam files using samtools^104^. QC measures of aligned reads was generated using picard metrics (https://slowkow.github.io/picardmetrics) and the CollectRnaSeqMetrics tool from picard tools (http://broadinstitute.github.io/picard). We filtered the bam files for mitochondrial reads and Stringtie was applied to merge and assemble reference guided transcripts for gene level quantifications of raw counts, and transcripts per million (TPM)^105^. Gene expression levels in TPM were used for further analyses as this unit allows efficient comparison of gene expression levels between samples from differed studies. A pseudo-count of 1 was added to all TPM values and and converted to log2 scale before the heatmaps were plotted. Hierarchical clustering by euclidean distance, z-score calculation and plotting the heatmap was done using the R package ‘pheatmap’ (Kolde R, 2019, v1.0.12). Z-scores are calculated by subtracting the mean of TPM values in all samples for a gene and dividing by the standard deviation. Samples with only TPM>5 were considered for heatmap showing the GWAS genes.

## Extended Data Figures

**Extended Data Figure 1.**
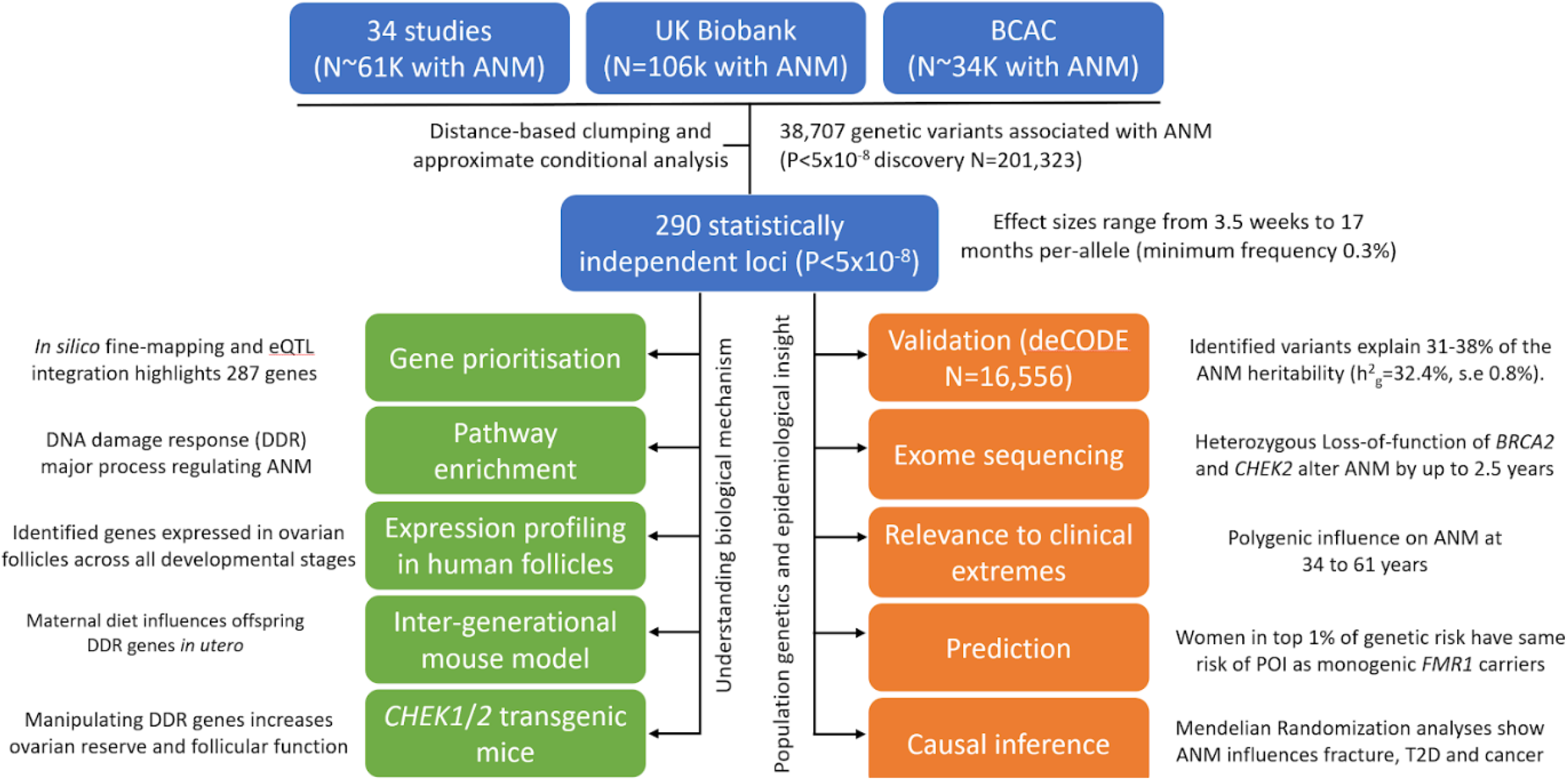
Overview of performed analyses. ANM, age at natural menopause; DDR, DNA damage response.

**Extended Data Figure 2.**
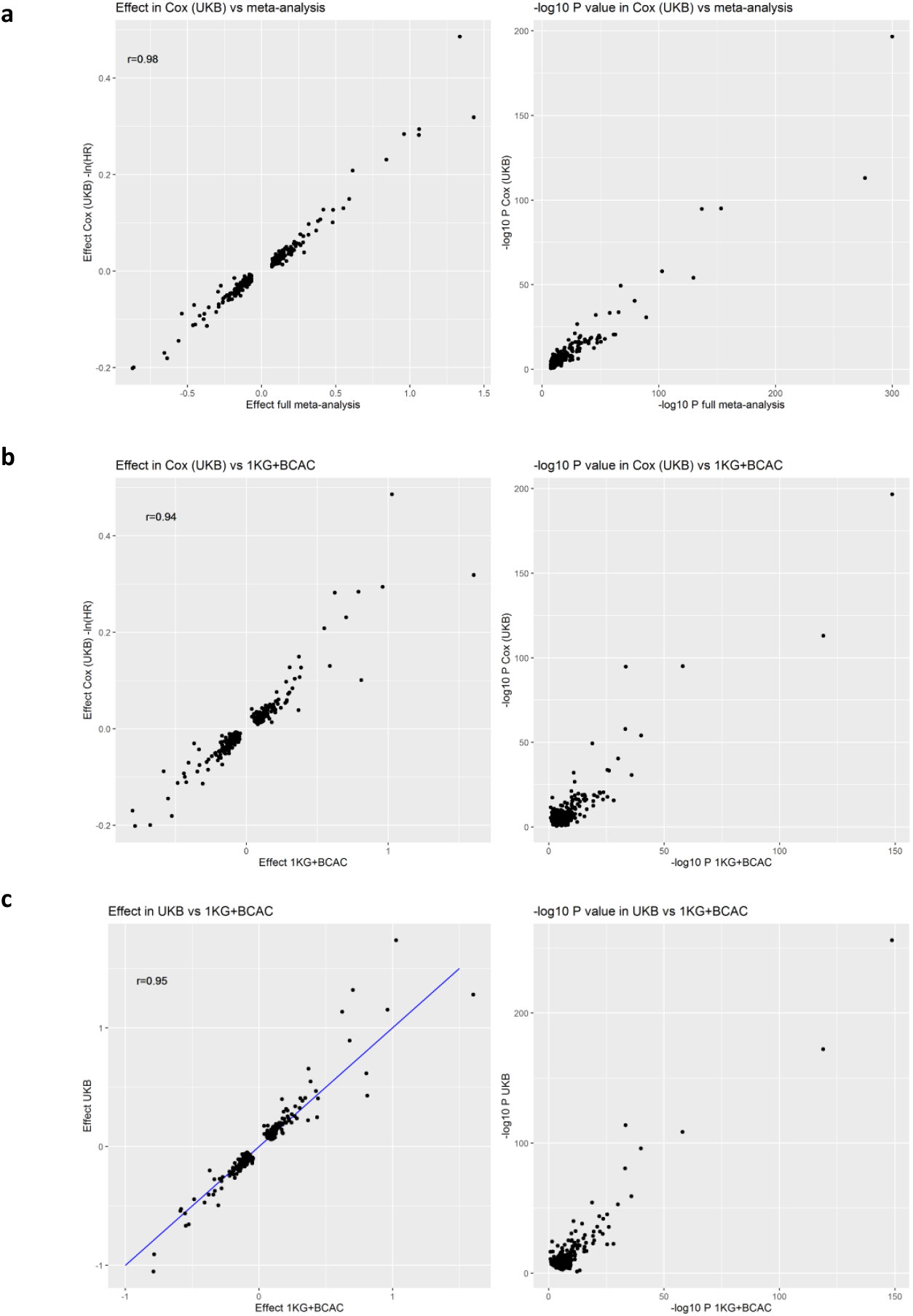
Consistency of effect estimates across analyses methods and strata. Comparison of effect estimates from: **a**, Cox proportional hazards regression in UK Biobank with linear regression effect estimates from the overall meta-analysis (“Effect full meta-analysis”); **b**, Cox proportional hazards regression in UK Biobank with linear regression effect estimates from the meta-analysis excluding UK Biobank (“Effect 1KG+BCAC”); **c**, linear regression in UK Biobank with linear regression effect estimates from the meta-analysis excluding UK Biobank (“Effect 1KG+BCAC”). HR, hazard ratio from Cox proportional hazards model; r, Pearson correlation coefficient. Note: *P* values < 1E-300 are shown as 1E-300.

**Extended Data Figure 3.**
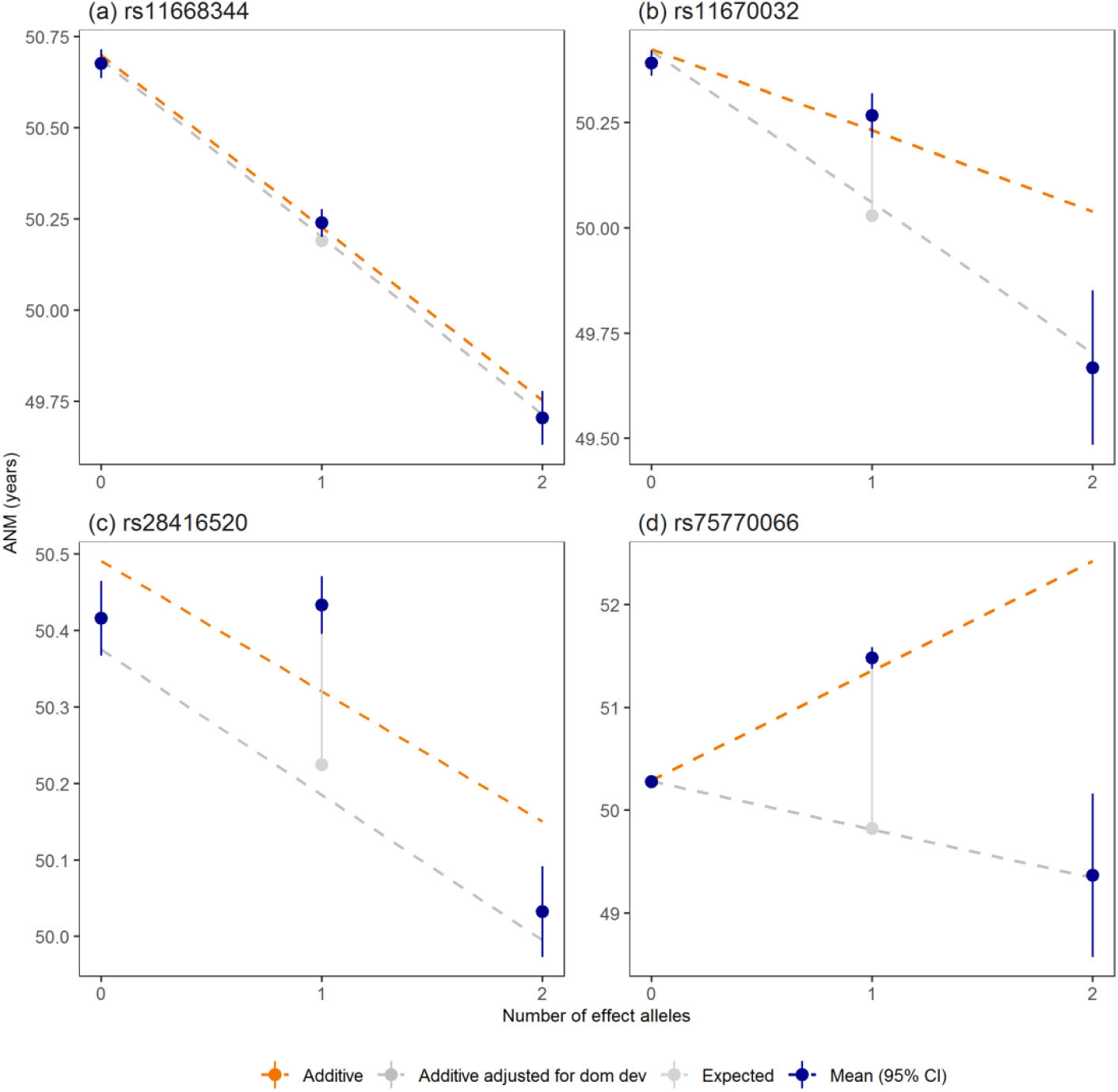
Genome-wide significant signals showing departure from an additive model. We tested the identified signals for departure from an additive allelic model. **a**, rs11668344 shows no deviation from an additive allelic model; **b**, rs11670032 and **c**, rs28416520 show deviation from the additive allelic model and a recessive effect; and **d**, rs75770066 shows a heterozygote effect. The mean and 95% confidence interval around the mean estimate are shown for each genotype. The expected mean ANM for the heterozygotes is the average of the mean ANM in the homozygote groups. The dashed orange line shows the effect estimate by genotype from linear regression based on an additive allelic model. Estimated ANM for each genotype was calculated as constant from regression model + number alleles × effect estimate from regression model. The dashed grey line indicates the additive effect estimate by genotype from a model adjusting for the dominance deviation effect of the heterozygote group (solid grey line). All regression models were adjusted for centre, genotyping chip and genetic principal components. ANM, age at natural menopause; dom dev, dominance deviation.

**Extended Data Figure 4.**
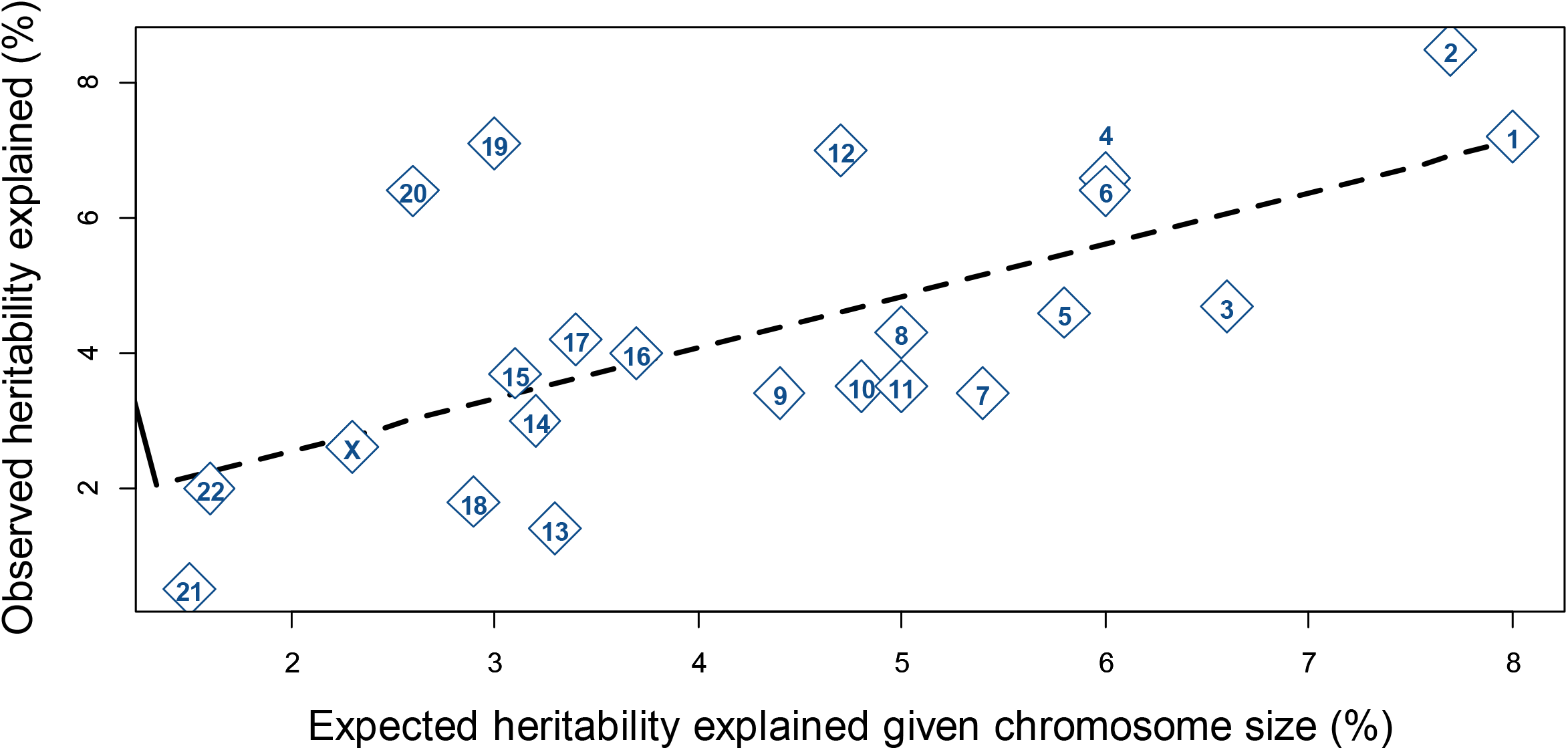
Distribution of estimated heritability across chromosomes, showing the mean ratio across all chromosomes.

**Extended Data Figure 5.**
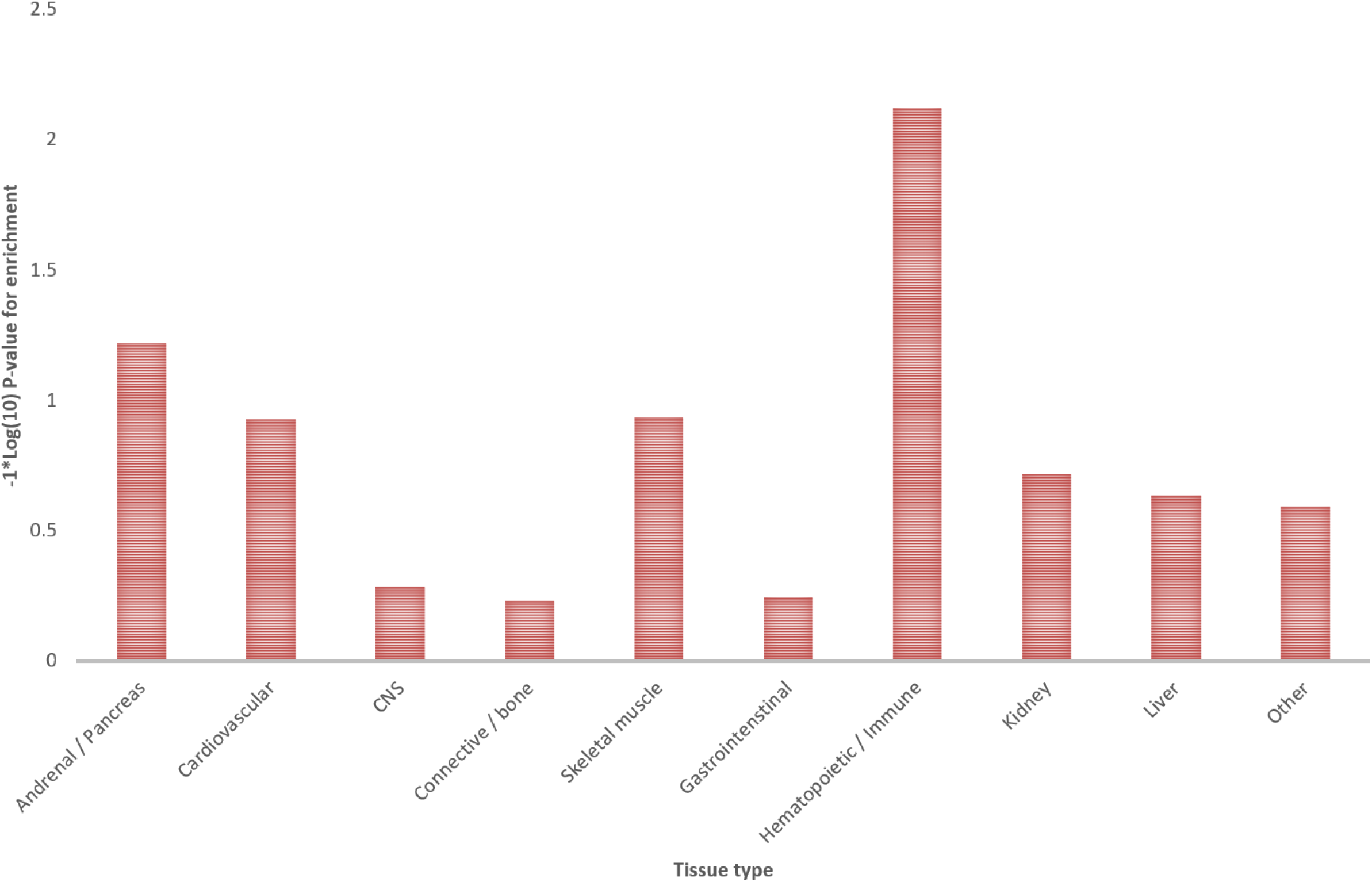
Tissue enrichment analyses performed using LDSC-SEG.

**Extended Data Figure 6.**
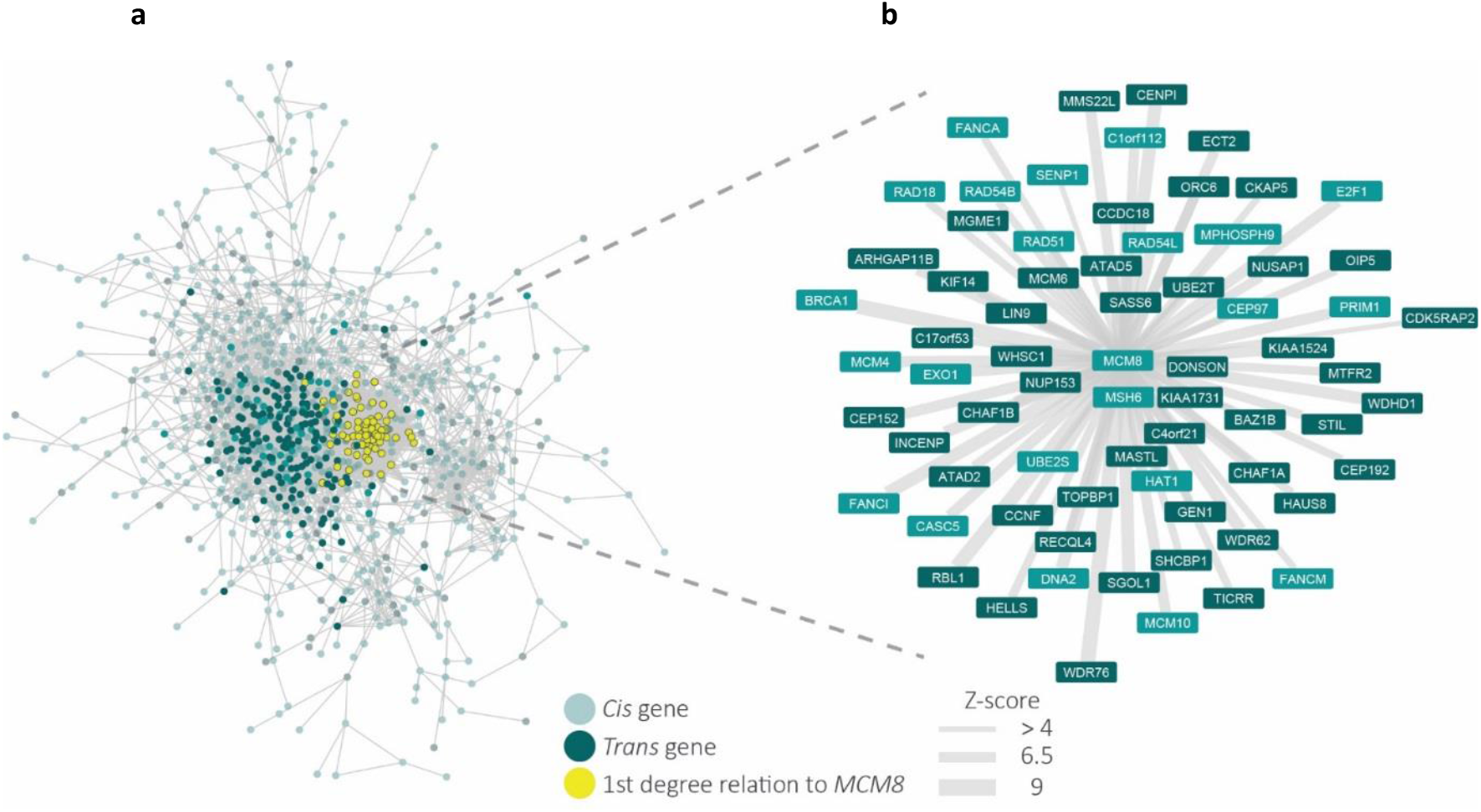
Gene co-regulation networks for age at menopause genes with those co-regulated with *MCM8* highlighted. **a**, Gene co-regulation network for genes relating to age at menopause. Nodes indicate genes that either in a *cis* region from the GWAS or have been prioritized by Downstreamer, edges indicate a co-regulation relationship with a Z-score >4. Co-regulation is defined as the Pearson correlation between genes in a scaled eigenvector matrix derived from a multi-tissue gene network [Deelen et al, Nat. Commun. 2019]. *Cis* genes are defined as genes that are within +/-300kb of a GWAS top hit for age at menopause. *Trans* genes are defined as having been prioritized by Downstreamer’s co-regulation analysis and are not within +/-300kb of a GWAS top hit. Downstreamer prioritizes genes by associating the gene p-value profile of the GWAS (calculated using PASCAL [Lamparter et al, PLOS Comput. Biol. 2016]) to the co-regulation profile of each protein coding gene. Only genes where this association passes Bonferroni significance are shown as trans genes. Colours of nodes indicate the following: Teal indicates *Cis* genes, Dark Teal indicates *Trans* genes and Yellow indicates genes with a 1st degree relation to *MCM8*. **b**, Gene co-regulation network showing the genes that have a first degree relationship with *MCM8* with a Z-score >4. Width of the edge indicates the Z-score of the co-regulation relationship. Colours indicate the same as in **a**, with the exception of Yellow, as all genes indicated have a 1st degree relation to *MCM8*.

**Extended Data Figure 7.**
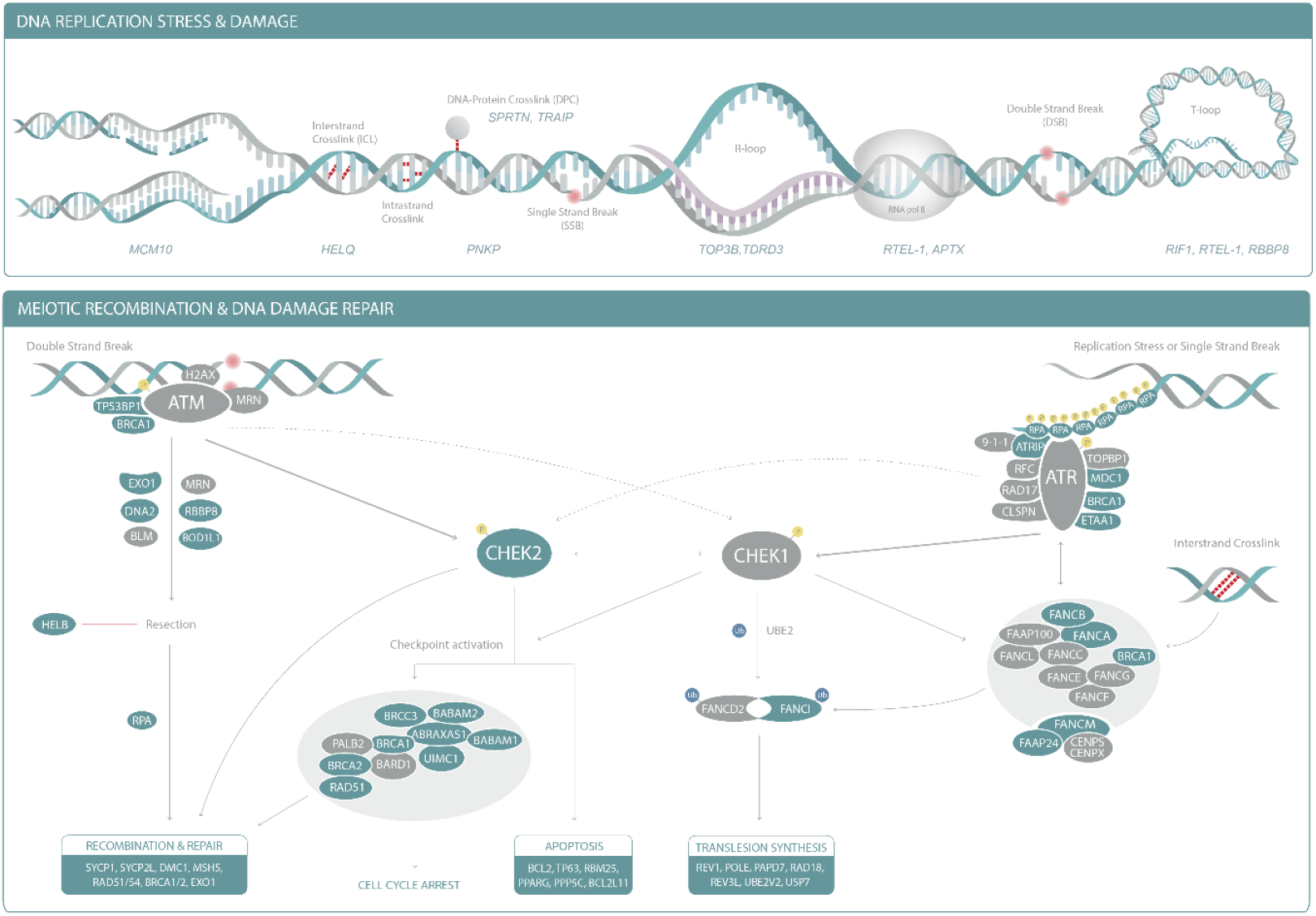
DNA damage response and repair pathways implicated in reproductive ageing in humans. Genes within 300kb of age at natural menopause (ANM) signals are listed in the top panel and are shaded in blue in the bottom panel. A full list of genes involved in DNA damage response and apoptosis annotated with genome-wide signals for ANM is provided in Supplementary Table 21. MRN, *MRN-MRE11-RAD50-NBS1* complex; RPA, Replication Protein A including a subunit encoded by *RPA1;* RFC, Replication Factor C including a subunit encoded by *RFC1*. 9-1-1, *RAD9-HUS1-RAD1* complex.

**Extended Data Figure 8.**
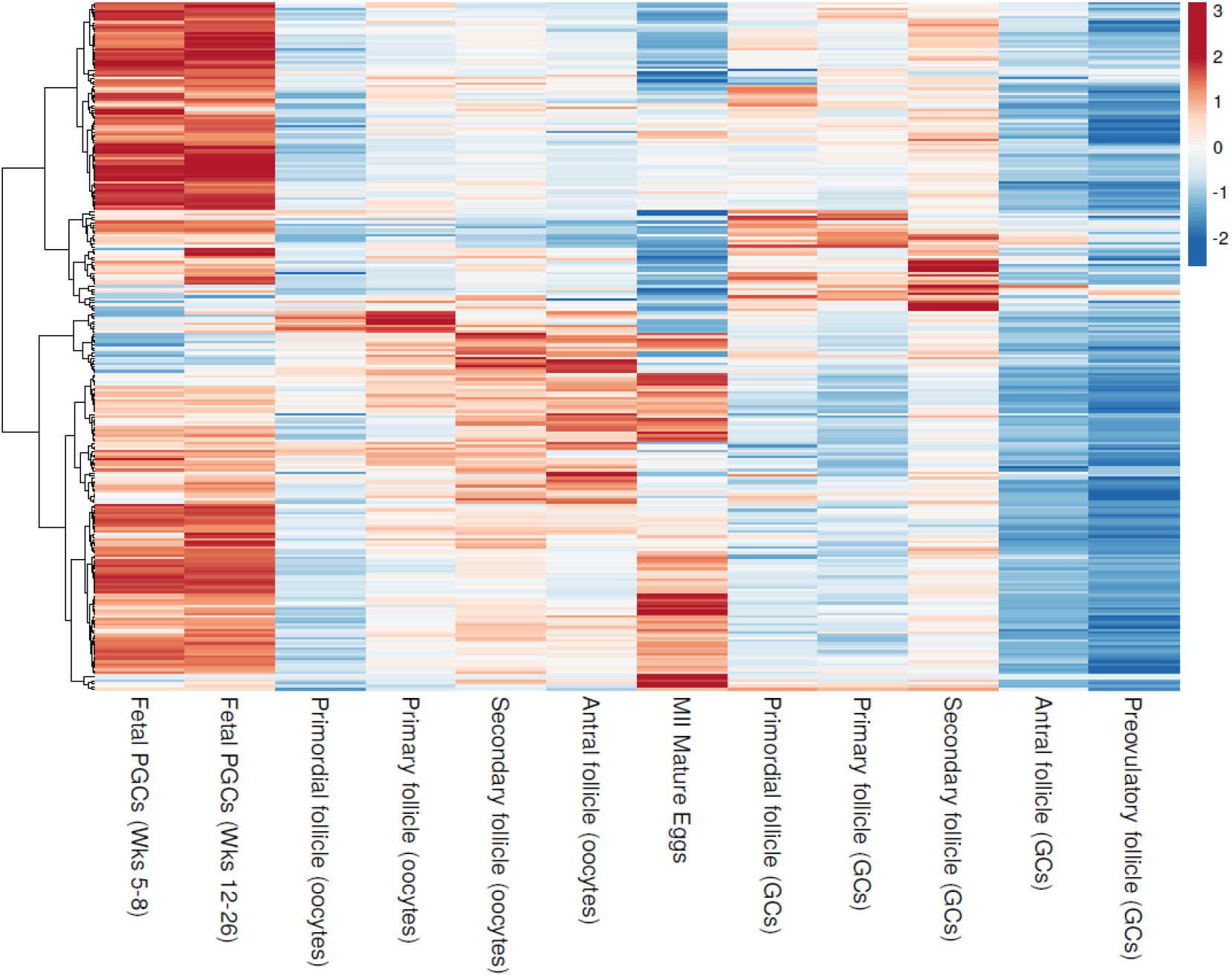
Cluster plot of expression of candidate genes identified from the genome-wide analyses in germ cells across different developmental stages. Gene expression was measured in human fetal primordial germ cells (Li et al 2017 and Zhang et al 2018), and oocytes and granulosa cells in adult follicles (dataset generated in this study). GC, granulosa cell; MII, meiosis II; PGC, primordial germ cell; Wks, weeks

**Extended Data Figure 9.**
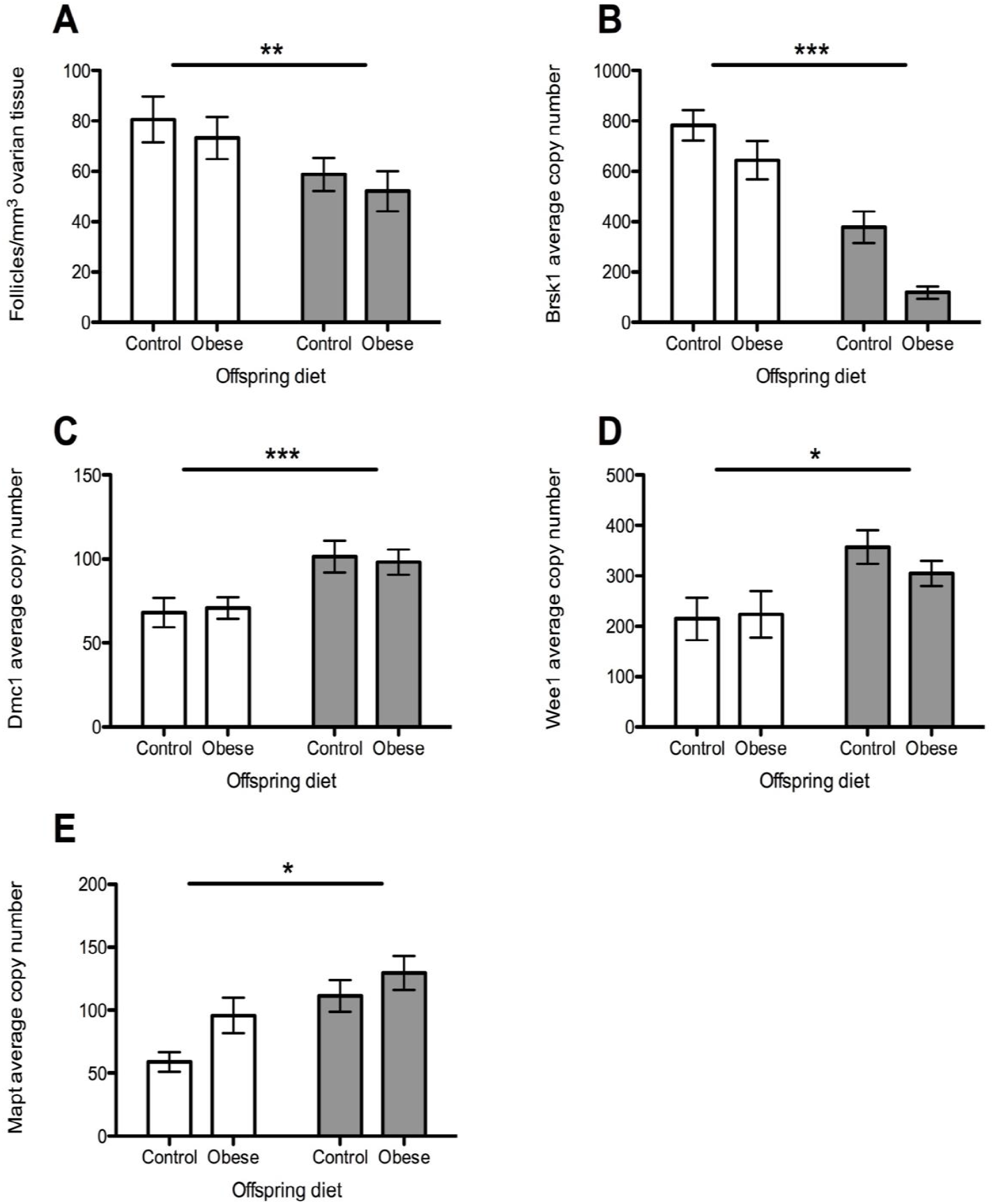
Relationship between decreased ovarian reserve and gene expression. Open bar groups: control maternal diet, normal ovarian reserve. Grey bar groups: obesogenic maternal diet, reduced ovarian reserve. **a**, Ovarian follicular reserve in young adulthood in wild-type mice. Total follicles/mm^3^ ovarian tissue at 12 weeks. **b**, *Brsk1* expression in the same animals, measured using qrtPCR and expressed as average copy number. **c**, *Dmc1* expression in the same animals, measured using qrtPCR and expressed as average copy number. **d**, *Wee1* expression in the same animals, measured using qrtPCR and expressed as average copy number. **e**, *Mapt* expression in the same animals, measured using qrtPCR and expressed as average copy number. Data are represented as means ± standard error. *, *P*<0.05; **, *P*<0.01; ***, *P*<0.001.

**Extended Data Figure 10.**
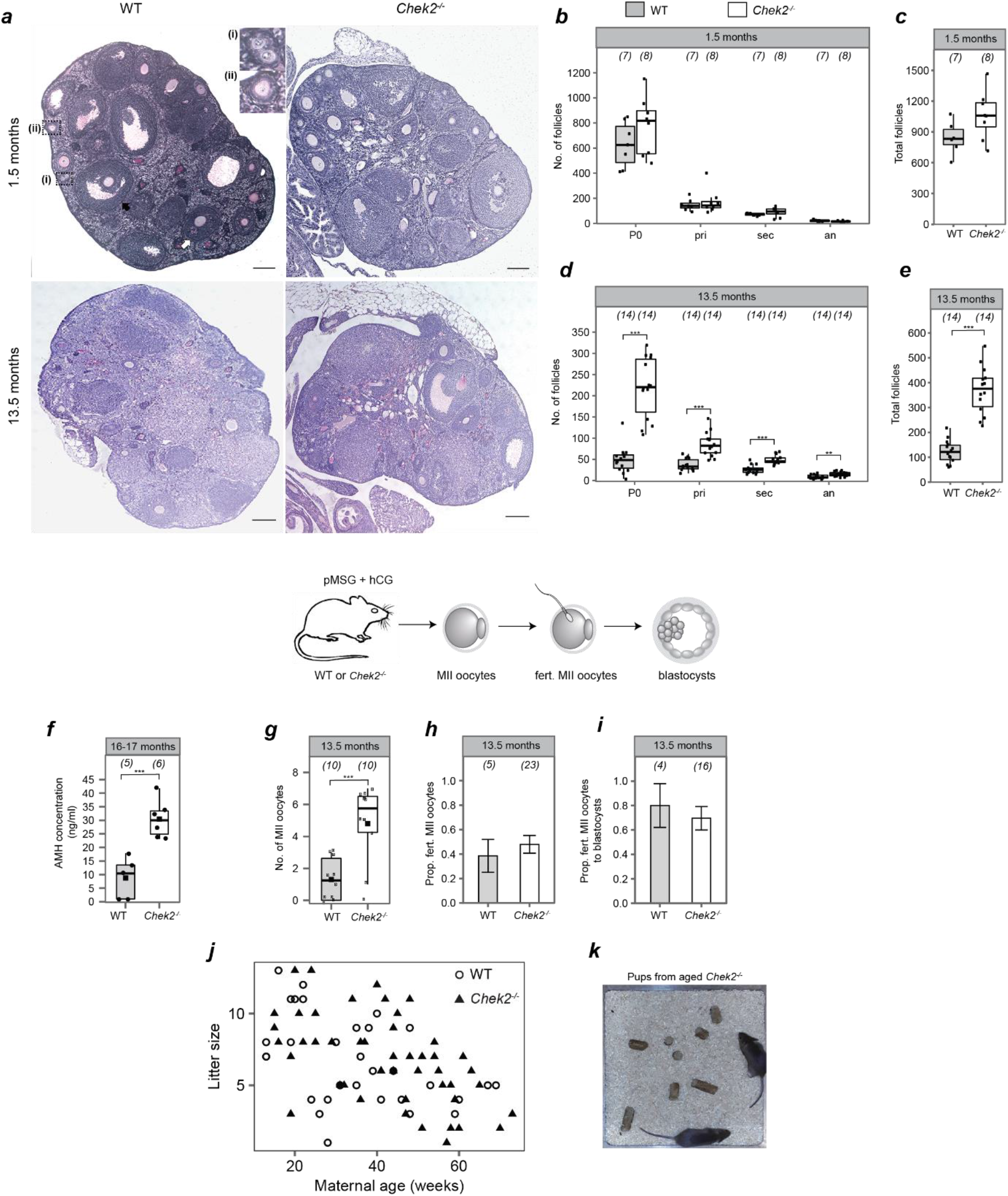
*Chek2* deletion increases reproductive lifespan in mouse. **a**, Representative images of ovarian sections of 1.5-and 13.5-month-old wild type (WT) and *Chek2*^*-/-*^ mice stained with PAS-Hematoxylin. Primordial follicles (inset (i)), primary follicles (inset (ii)), secondary follicle (white arrow) and antral follicle (black arrow) are shown. Scale bar represents 200 µm. **b-e**, Quantification of the number of follicles (by class and total) present in WT and *Chek2*^*-/-*^ mice ovaries: **b, c**, 1.5-month-old; **d, e**, 13.5-month-old. The numbers in parentheses correspond to the total number of ovaries analysed. **f**, Serum AMH (ng/ml) in 16-17 months old Chek2-/- mice. The numbers in parentheses correspond to the number of mice assessed. **g-i**, Diagram illustrates the gonadotrophin stimulation of 13.5-month old females. Numbers in parentheses show: **g**, the number of MII oocytes retrieved per female; **h**, the number of MII oocytes fertilized; and **i**, the number of fertilized oocytes assessed for blastocyst development. **j**, Litter size of WT and *Chek2*^*-/-*^ females throughout the reproductive life span. Litter sizes from 9 WT and 5 *Chek2*^−/-^ females are shown. Breeding cages contained one male and one female. Generalized linear model analysis showed maternal age effect, but no effect on genotype on litter sizes. **k**, Image of healthy pups born to 13 month-old *Chek2*^*-/-*^ females. **b-i**, Two sample *t* and Fisher’s exact tests were used to compare WT and *Chek2*^*-/-*^ for statistical significance: *, *P*<0.05; **, *P*<0.025; ***, *P*<0.001. Error bars indicate standard error of mean. an, antral follicle; hCG, human chorionic gonadotrophin; pMSG, pregnant mare serum gonadotrophin; pri, primary follicle; P0, primordial follicle; sec, secondary follicle; WT=wildtype.

**Extended Data Figure 11.**
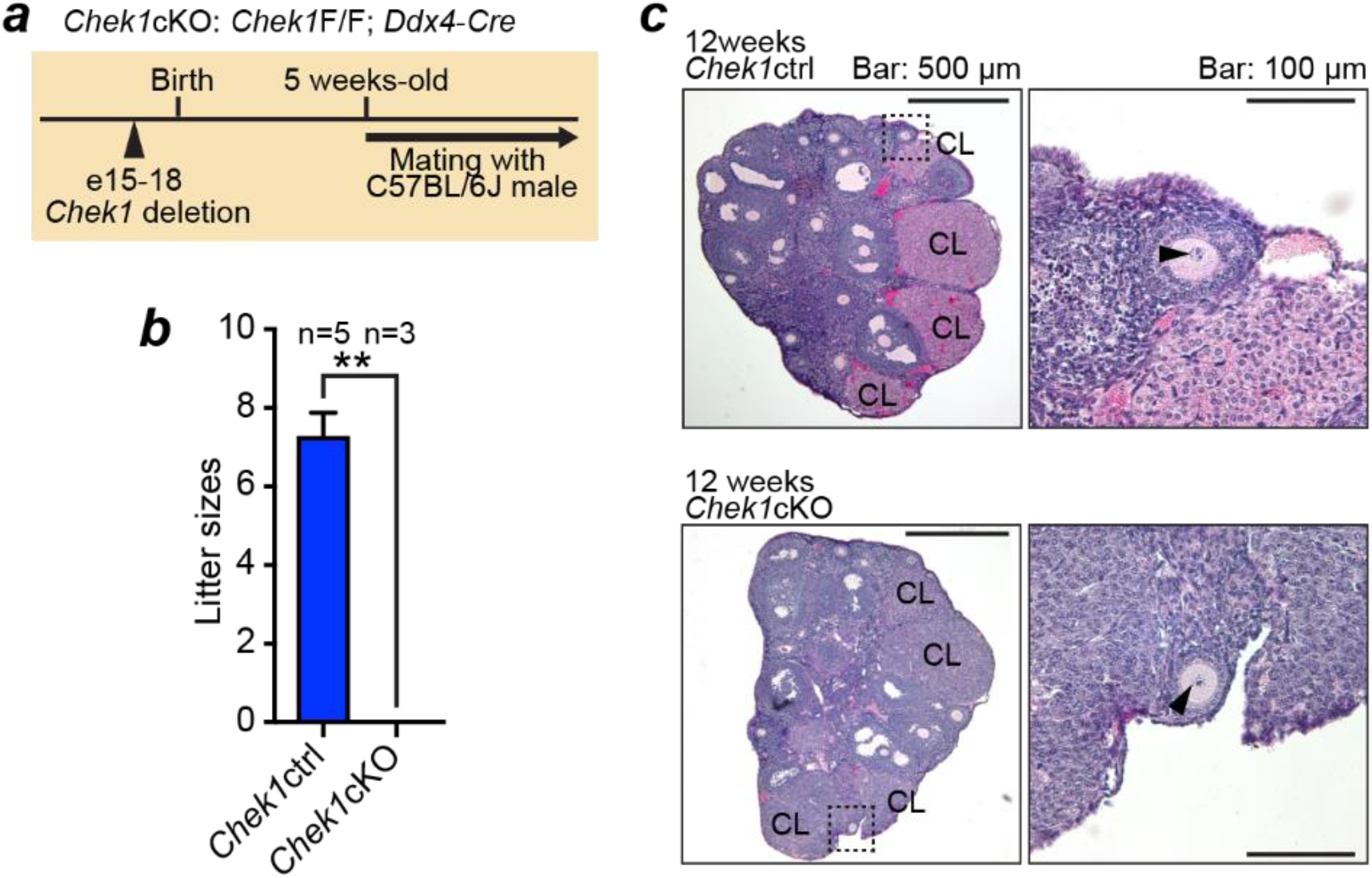
Conditional knockout *Chek1* females are infertile. **a**, Schematic of the conditional-knockout mouse model of *Chek1* (*Chek1-cKO*) in the female germline using the *Ddx4*-Cre transgene. **b**, To assess the fertility of *Chek1*cKO, three independent females older than 5 weeks age were mated with C57BL/6J males. Five independent littermate females (F/+, Tg-/Tg-; F/F, Tg-/Tg-; or F/+, Tg+/Tg-) were used as *Chek1* controls (ctrl). While *Chek1*ctrl females delivered normally, *Chek1*cKO females delivered no litters (**, Mann Whitney test *P*=0.0179). Thus, these results indicate that *CHEK1* is essential in the female germline. **c**, In the sections stained with haematoxylin and eosin, we found follicles, corpora lutea (CL) and oocytes which contain nuclear structures (indicated with arrowheads in the magnified right hand panel). These findings suggest that estrus cycles and ovulation followed by corpus luteum formation are independent from *Chek1* disruption in oocytes *in vivo*.

**Extended Data Figure 12.**
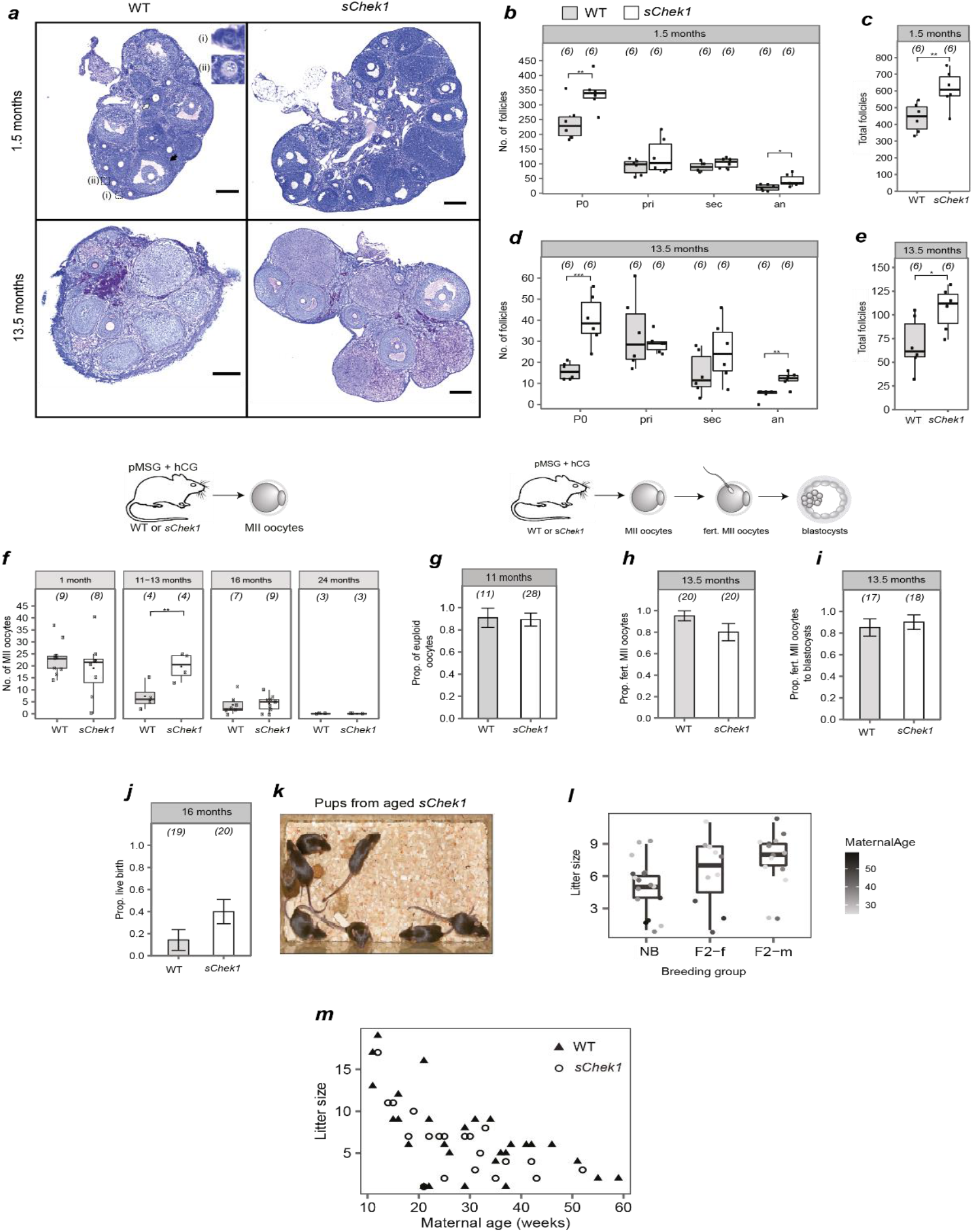
Extended reproductive lifespan in females carrying an extra copy of *Chek1 (sChek1)*. **a**, Representative images of ovarian sections of 1.5 and 13.5-month-old wild type (WT) and *sChek1* mice stained with PAS-hematoxylin. Primordial follicles (inset (i)), primary follicles (inset (ii)), secondary follicle (white arrow) and antral follicle (black arrow) are shown. Scale bar: 200 µm. **b-e**, Quantification of the number of follicles (by class and total) present in WT and *sChek1* litter mates: **b, c**, 1.5-month-old; **d, e**, 13.5-month-old. The numbers in parentheses correspond to the total number of ovaries analysed. **f-i**, MII oocytes retrieved in response to pMSG and hCG, proportion of euploid oocytes, proportion fertilized and proportion developed to blastocysts at different ages of WT and *sChek1* mice. Numbers in parentheses show: **f**, the number of MII oocytes retrieved per female; **g**, the number of oocytes assessed for aneuploidy; **h**, the number of MII oocytes fertilized; and **i**, the number of fertilized oocytes assessed for blastocyst development. **j**, Proportion of live births relative to transferred embryos from *in vitro* fertilized oocytes from aged mice (16 months), the numbers in parenthesis show the embryos transferred. **k**, Photo of healthy pups born to 16-month old *sChek1* females after IVF. **l**, Litter sizes from F2 females or males from aged *sChek1* females after IVF treatment in **j**, compared to females of equivalent ages that were naturally breeding. Note that for natural breeding there were two females and one male per breeding cage, whereas F2 cages contained a single male and one female. Therefore, litter sizes are an underestimate for the IVF-conceived pups. **m**, Litter sizes of *sChek1* and WT females throughout their reproductive life span. Data are from six breeding cages, three for each genotype. Each breeding cage contained one WT male and two females that were either WT or *sChek1*. Generalized linear model analysis showed maternal age effect, but no effect on genotype on litter sizes. **b-j, l**, Two sample *t* and Fisher’s exact tests were used to compare WT and s*Chek1* for statistical significance: *, *P*<0.05; **, *P*<0.025; ***, *P*<0.001. Error bars indicate standard error of mean. an=antral follicle; hCG= human chorionic gonadotrophin; IVF=*in vitro* fertilization; NB=naturally breeding; pMSG=pregnant mare serum gonadotrophin; pri=primary follicle; P0=primordial follicle; sec=secondary follicle; WT=wildtype.

