## Supplementary Information for "Genetic insights into the biological mechanisms governing human ovarian ageing"

**Contents:**

**1) Acknowledgments and sources of funding**

**2) Individual study disclosures**

**Acknowledgments and sources of funding:**

**1) Studies included in the genome-wide association analyses:**

| **Study acronym** | **Full study name** | **Acknowledgments and sources of funding** |
| --- | --- | --- |
| AGES | Age, Gene/Environment Susceptibility-Reykjavik Study | This study has been funded by NIH contracts N01-AG-1-2100 and 271201200022C, the NIA Intramural Research Program, Hjartavernd (the Icelandic Heart Association), and the Althingi (the Icelandic Parliament). The study is approved by the Icelandic National Bioethics Committee, VSN: 00-063. The researchers are indebted to the participants for their willingness to participate in the study. |
| ALSPAC | Avon Longitudinal Study of Parents and Children | We are extremely grateful to all the families who took part in this study, the midwives for their help in recruiting them, and the whole ALSPAC team, which includes interviewers, computer and laboratory technicians, clerical workers, research scientists, volunteers, managers, receptionists and nurses.  The UK Medical Research Council and the Wellcome Trust (Grant ref: 102215/2/13/2) and the University of Bristol provide core support for ALSPAC. GWAS data for ALSPAC mothers was funded by the Wellcome Trust (WT088806) and phenotype data by the British Heart Foundation (SP/07/008/24066), Wellcome Trust (WT092830M) and MRC (G1001357). GWAS data for ALSPAC offspring was generated by Sample Logistics and Genotyping Facilities at the Wellcome Trust Sanger Institute and LabCorp (Laboratory Corporation of America) using support from 23andMe. DAL, NJT, SMR and GDS work in a Unit that receives support from the University of Bristol and MRC (MC_UU_12013/1; MC_UU_12013/2 and MC_UU_12013/5). |
| ARIC | Atherosclerosis Risk in Communities Study | The Atherosclerosis Risk in Communities study has been funded in whole or in part with Federal funds from the National Heart, Lung, and Blood Institute, National Institutes of Health, Department of Health and Human Services, under Contract nos.  (HHSN268201700001I, HHSN268201700002I, HHSN268201700003I,  HHSN268201700005I, HHSN268201700004I).  The authors thank the staff and participants of the ARIC study for their important  contributions.  The Atherosclerosis Risk in Communities study has been funded in whole or in part with Federal funds from the National Heart, Lung, and Blood Institute, National Institutes of Health, Department of Health and Human Services (contract numbers HHSN268201700001I, HHSN268201700002I,  HHSN268201700003I, HHSN268201700004I and HHSN268201700005I), R01HL087641, R01HL086694; National Human Genome Research Institute contract U01HG004402; and National Institutes of Health contract HHSN268200625226C. The authors thank the staff and participants of the ARIC study for their important contributions. Infrastructure was partly supported by Grant Number UL1RR025005, a component of the National Institutes of Health and NIH Roadmap for Medical Research. |
| BCAC/ iCOGs | Breast Cancer Association Consortium/ iCOGs | See separate section below for details of individual studies. |
| CARL | INGI-CARLANTINO | We would like to thank all the participants in the study for their contribution and support  The study was supported through the Italian Ministry of Health |
| CHS | Cardiovascular Health Study | This CHS research was supported by NHLBI contracts HHSN268201200036C, HHSN268200800007C, HHSN268201800001C, N01HC55222, N01HC85079, N01HC85080, N01HC85081, N01HC85082, N01HC85083, N01HC85086; and NHLBI grants U01HL080295, R01HL087652, R01HL105756, R01HL103612, R01HL120393, and U01HL130114 with additional contribution from the National Institute of Neurological Disorders and Stroke (NINDS). Additional support was provided through R01AG023629 from the National Institute on Aging (NIA). A full list of principal CHS investigators and institutions can be found at CHS-NHLBI.org.  The provision of genotyping data was supported in part by the National Center for Advancing Translational Sciences, CTSI grant UL1TR001881, and the National Institute of Diabetes and Digestive and Kidney Disease Diabetes Research Center (DRC) grant DK063491 to the Southern California Diabetes Endocrinology Research Center.  The content is solely the responsibility of the authors and does not necessarily represent the official views of the National Institutes of Health. |
| CILENTO | CILENTO | We thank the populations of Cilento for their participation in the study.  This work was supported by grants from the Italian Ministry of Universities (FIRB-RBNE08NKH7, INTEROMICS Flagship Project), the Assessorato Ricerca Regione Campania, the Fondazione con il SUD (2011-PDR-13) and the Istituto Banco di Napoli - Fondazione to MC. |
| COLAUS | CoLaus (Etude Cohorte Lausannoise) | The CoLaus study was and is supported by research grants from GlaxoSmithKline, the Faculty of Biology and Medicine of Lausanne, and the Swiss National Science Foundation (grants 33CSCO-122661, 33CS30-139468, 33CS30-148401 and 32473B-182210). |
| CROATIA Korcula | CROATIA Korcula | We would like to acknowledge the contributions of the recruitment team in Korcula, the administrative teams in Croatia and Edinburgh and the people of Korcula. The SNP genotyping for the KORCULA cohort was performed in Helmholtz Zentrum München, Neuherberg, Germany.  Medical Research Council UK and the Ministry of Science, Education and Sport in the Republic of Croatia (number 108-1080315-0302). |
| CROATIA Vis | CROATIA Vis | We would like to acknowledge the staff of several institutions in Croatia that supported the field work, including but not limited to The University of Split and Zagreb Medical Schools, Institute for Anthropological Research in Zagreb and Croatian Institute for Public Health.  Medical Research Council UK and the Ministry of Science, Education and Sport in the Republic of Croatia (number 108-1080315-0302). |
| EGCUT-370 and OmniX | Estonian Genome Center, University of Tartu | This study was supported by EU H2020 grants 692145, 676550, 654248, Estonian Research Council Grant IUT20-60, NIASC, and EU through the European Regional Development Fund (Project No. 2014-2020.4.01.15-0012 GENTRANSMED). |
| EPIC-Norfolk | The EPIC-Norfolk Study |  |
| FHS | Framingham Heart Study | The authors thank the Framingham Heart Study participants and staff.  The Framingham Heart Study phenotype-genotype analyses were supported by NIA R01AG29451 JMM, KLL). The Framingham Heart Study of the National Heart Lung and Blood Institute of the National Institutes of Health and Boston University School of Medicine was supported by the National Heart, Lung and Blood Institute's Framingham Heart Study Contract No. N01-HC-25195 and its contract with Affymetrix, Inc for genotyping services (Contract No. N02-HL-6-4278). Analyses reflect intellectual input and resource development from the Framingham Heart Study investigators participating in the SNP Health Association Resource (SHARe) project. A portion of this research was conducted using the Linux Cluster for Genetic Analysis (LinGA-II) funded by the Robert Dawson Evans Endowment of the Department of Medicine at Boston University School of Medicine and Boston Medical Center.  Framingham Heart Study Contract No. N01-HC-25195, HHSN268201500001  The authors are pleased to acknowledge that the computational work reported on in this paper was performed on the Shared Computing Cluster which is administered by Boston University’s Research Computing Services. URL: www.bu.edu/tech/support/research/. The authors thank the participants for their dedication to the study. |
| FVG | INGI- FRIULI VENEZIA GIULIA | We would like to thank all the participants in the study for their contribution and support  The study was supported by Regione FVG (L.26.2008) and Italian Ministry of Health |
| GS | Generation Scotland: Scottish Family Health Study | We are grateful to all the families who took part, the general practitioners and the Scottish School of Primary Care for their help in recruiting them, and the whole Generation Scotland team, which includes interviewers, computer and laboratory technicians, clerical workers, research scientists, volunteers, managers, receptionists, healthcare assistants and nurses. Genotyping was performed at the Wellcome Trust Clinical Research Facility Genetics Core at Western General Hospital, Edinburgh, UK.  Scottish Executive Health Department, Chief Scientist Office, grant number CZD/16/6. Exome array genotyping for GS:SFHS was funded by the Medical Research Council UK |
| HANDLS |  |  |
| Health ABC | The Health, Aging, and Body Composition Study | This study utilized the high-performance computational capabilities of the Biowulf Linux cluster at the National Institutes of Health, Bethesda, Md. (http://biowulf.nih.gov).  The Health ABC Study was supported by NIA contracts N01AG62101, N01AG62103, and N01AG62106 and, in part, by the NIA Intramural Research Program. The genome-wide association study was funded by NIA grant 1R01AG032098-01A1 to Wake Forest University Health Sciences and genotyping services were provided by the Center for Inherited Disease Research (CIDR). CIDR is fully funded through a federal contract from the National Institutes of Health to The Johns Hopkins University, contract number HHSN268200782096C. |
| HRS | Health and Retirement Study | HRS is supported by the National Institute on Aging (NIA U01AG009740). The genotyping was funded separately by the National Institute on Aging (RC2 AG036495, RC4 AG039029). Our genotyping was conducted by the NIH Center for Inherited Disease Research (CIDR) at Johns Hopkins University. Genotyping quality control and final preparation of the data were performed by the Genetics Coordinating Center at the University of Washington. |
| INCHIANTI | Invecchiare in Chianti | The InCHIANTI study baseline (1998-2000) was supported as a "targeted project" (ICS110.1/RF97.71) by the Italian Ministry of Health and in part by the U.S. National Institute on Aging (Contracts: 263 MD 9164 and 263 MD 821336). |
| InterAct cases and cohort | European Prospective Investigation into Cancer & Nutrition - InterAct |  |
| KORA F3 / KORA F4 | KORA F3 and F4: Cooperative Health Research in the Augsburg Region, Follow up for Study S3 and S4 | The KORA study was initiated and financed by the Helmholtz Zentrum München – German Research Center for Environmental Health, which is funded by the German Federal Ministry of Education and Research (BMBF) and by the State of Bavaria. The KORA-Study Group consists of A. Peters (speaker), H. Schulz, R. Holle, R. Leidl, C. Meisinger, K. Strauch, and their co-workers, who are responsible for the design and conduct of the KORA studies. Furthermore, KORA research was supported within the Munich Center of Health Sciences (MC-Health), Ludwig-Maximilians-Universität, as part of LMUinnovativ. The funders had no role in study design, data collection and analysis, decision to publish, or preparation of the manuscript. We thank all study participants and the study staff. |
| LifeLines | The LifeLines Cohort Study | The authors wish to acknowledge Behrooz Z. Alizadeh, Annemieke Boesjes, Marcel Bruinenberg, Noortje Festen, Pim van der Harst, Ilja Nolte, Lude Franke, Mitra Valimohammadi for their help in creating the GWAS database, and Rob Bieringa, Joost Keers, René Oostergo, Rosalie Visser, Judith Vonk for their work related to data-collection and validation. The authors are grateful to the study participants, the staff from the LifeLines Cohort Study and the contributing research centers delivering data to LifeLines and the participating general practitioners and pharmacists.  The Lifelines Biobank initiative has been made possible by subsidy from the Dutch Ministry of Health, Welfare and Sport, the Dutch Ministry of Economic Affairs, the University Medical Center Groningen (UMCG the Netherlands), University Groningen and the Northern Provinces of the Netherlands.  The Lifelines Cohort Study was supported by the Netherlands Organization for Scientific Research (NWO) [grant 175.010.2007.006]; the Economic Structure Enhancing Fund (FES) of the Dutch government; the Ministry of Economic Affairs; the Ministry of Education, Culture and Science; the Ministry for Health, Welfare and Sports; the Northern Netherlands Collaboration of Provinces (SNN); the Province of Groningen; University Medical Center Groningen; the University of Groningen; the Dutch Kidney Foundation; and the Dutch Diabetes Research Foundation. The Lifelines Cohort Study is supported by the National Consortium for Healthy Ageing and the BioSHaRE-EU consortium (KP7, project reference 261433). The funders had no role in study design, data collection and analysis, decision to publish, or preparation of the manuscript. |
| NEO | Netherlands Epidemiology of Obesity | The authors of the NEO study thank all individuals who participated in the Netherlands Epidemiology in Obesity study, all participating general practitioners for inviting eligible participants and all research nurses for collection of the data. We thank the NEO study group, Pat van Beelen, Petra Noordijk and Ingeborg de Jonge for the coordination, lab and data management of the NEO study.  The genotyping in the NEO study was supported by the Centre National de Génotypage (Paris, France), headed by Jean-Francois Deleuze. The NEO study is supported by the participating Departments, the Division and the Board of Directors of the Leiden University Medical Center, and by the Leiden University, Research Profile Area Vascular and Regenerative Medicine. Dennis Mook-Kanamori is supported by Dutch Science Organization (ZonMW-VENI Grant 916.14.023). |
| NHS Affymetrix / NHS Illumina / NHS Omni-Express | The Nurses’ Health Study (NHS) | The NHS GWAS were supported by grants from the National Institutes of Health [NCI (CA40356, CA087969, CA055075, CA98233, U01 CA137088, R01 CA059045, R01 CA137178, R01 CA082838, R01 CA131332), NIDDK (DK058845, DK070756), NHGRI (HG004399, HG004728), NHLBI (HL35464), NIAMS (R01 AR056291)]. We would like to thank the participants and staff of the NHS and NHSII for their valuable contributions as well as the following state cancer registries for their help: AL, AZ, AR, CA, CO, CT, DE, FL, GA, ID, IL, IN, IA, KY, LA, ME, MD, MA, MI, NE, NH, NJ, NY, NC, ND, OH, OK, OR, PA, RI, SC, TN, TX, VA, WA, WY. The authors assume full responsibility for analyses and interpretation of these data. |
| NTR | The Netherlands Twin Register | The Netherland Twin Register: would like to thank all study participants for their contributions to our scientific efforts, the SURF SARA institute for computational resources and the Avera institute of Human Genetics for genotyping of samples.  Funding was obtained from the Netherlands Organization for Scientific Research (NWO) and The Netherlands Organization for Health Research and Development (ZonMW) grants 904-61-090, 985-10-002, 912-10-020, 904-61-193,480-04-004, 463-06-001, 451-04-034, 400-05-717, Addiction-31160008, 016-115-035, 481-08-011, 400-07-080, 056-32-010, Middelgroot-911-09-032, OCW_NWO Gravity program –024.001.003, NWO-Groot 480-15-001/674, Center for Medical Systems Biology (CSMB, NWO Genomics), NBIC/BioAssist/RK(2008.024), Biobanking and Biomolecular Resources Research Infrastructure (BBMRI –NL, 184.021.007 and 184.033.111), X-Omics 184-034-019; Spinozapremie (NWO- 56-464-14192), KNAW Academy Professor Award (PAH/6635) and University Research Fellow grant (URF) to DIB; Amsterdam Public Health research institute (former EMGO+) , Neuroscience Amsterdam research institute (former NCA); Amsterdam Research & Development (AR&D) research institute; the European Community's Fifth and Seventh Framework Program (FP5- LIFE QUALITY-CT-2002-2006, FP7- HEALTH-F4-2007-2013, grant 01254: GenomEUtwin, grant 01413: ENGAGE and grant 602768: ACTION); the European Research Council (ERC Starting 284167, ERC Consolidator 771057, ERC Advanced 230374), Rutgers University Cell and DNA Repository (NIMH U24 MH068457-06), the National Institutes of Health (NIH, R01D0042157-01A1, R01MH58799-03, MH081802, DA018673, R01 DK092127-04, Grand Opportunity grants 1RC2 MH089951, and 1RC2 MH089995); the Avera Institute for Human Genetics, Sioux Falls, South Dakota (USA). Part of the genotyping and analyses were funded by the Genetic Association Information Network (GAIN) of the Foundation for the National Institutes of Health. Computing was supported by NWO through grant 2018/EW/00408559, BiG Grid, the Dutch e-Science Grid and SURFSARA. |
| ORCADES | Orkney Complex Disease Study | DNA extractions were performed at the Wellcome Trust Clinical Research Facility in Edinburgh. We would like to acknowledge the invaluable contributions of the research nurses in Orkney, the administrative team in Edinburgh and the people of Orkney.  ORCADES was supported by the Chief Scientist Office of the Scottish Government (CZB/4/276, CZB/4/710), the Royal Society, the MRC Human Genetics Unit, Arthritis Research UK and the European Union framework program 6 EUROSPAN project (contract no. LSHG-CT-2006-018947). |
| QIMR | QIMR Adult Cohort | We thank the participants and their families for contributing to this research. We also thank A Henders, B Usher, E Souzeau, A Kuot, A McMellon, MJ Wright, MJ Campbell, A Caracella, L Bowdler, S Smith, B Haddon, A Conciatore, D Smyth, H Beeby, O Zheng and B Chapman for their input into project management, databases, phenotype collection, and sample collection, processing and genotyping.  The QIMR cohort was supported by National Institutes of Health (NIH) Grants AA07535, AA07728, AA13320, AA13321, AA14041, AA11998, AA17688, DA012854, DA019951, AA010249, AA013320, AA013321, AA011998, AA017688, and DA027995; by Grants from the Australian National Health and Medical Research Council (NHMRC) (241944, 339462, 389927, 389875, 389891, 389892, 389938, 442915, 442981, 496739, 552485, 552498 and 1075175); by Grants from the Australian Research Council (ARC) (A7960034, A79906588, A79801419, DP0770096, DP0212016, and DP0343921); DRN (FT0991022, 613674) SEM (1103623) and GWM (619667) were supported by the ARC Future Fellowship and NHMRC Fellowship Schemes. |
| RSI / RSII / RSIII | Rotterdam Study I, 2 and 3 | The Rotterdam Study (PMID: 32367290) is funded by Erasmus Medical Center and Erasmus University, Rotterdam, Netherlands Organization for the Health Research and Development (ZonMw), the Research Institute for Diseases in the Elderly (RIDE), the Ministry of Education, Culture and Science, the Ministry for Health, Welfare and Sports, the European Commission (DG XII), and the Municipality of Rotterdam. The authors are grateful to the study participants, the staff from the Rotterdam Study and the participating general practitioners and pharmacists. The generation and management of GWAS genotype data for the Rotterdam Study (RS I, RS II, RS III) was executed by the Human Genotyping Facility of the Genetic Laboratory of the Department of Internal Medicine, Erasmus MC, Rotterdam, The Netherlands. The GWAS datasets are supported by the Netherlands Organisation of Scientific Research NWO Investments (nr. 175.010.2005.011, 911-03-012), the Genetic Laboratory of the Department of Internal Medicine, Erasmus MC, the Research Institute for Diseases in the Elderly (014-93-015; RIDE2), the Netherlands Genomics Initiative (NGI)/Netherlands Organisation for Scientific Research (NWO) Netherlands Consortium for Healthy Aging (NCHA), project nr. 050-060-810. We thank Pascal Arp, Mila Jhamai, Marijn Verkerk, Lizbeth Herrera and Marjolein Peters, MSc, and Carolina Medina-Gomez, MSc, for their help in creating the GWAS database, and Karol Estrada, PhD, Yurii Aulchenko, PhD, and Carolina Medina-Gomez, MSc, for the creation and analysis of imputed data.  The Rotterdam Study has been approved by the Medical Ethics Committee of the Erasmus MC (registration number MEC 02.1015) and by the Dutch Ministry of Health, Welfare and Sport (Population Screening Act WBO, license number 1071272-159521-PG). The Rotterdam Study Personal Registration Data collection is filed with the Erasmus MC Data Protection Officer under registration number EMC1712001. The Rotterdam Study has been entered into the Netherlands National Trial Register (NTR; www.trialregister.nl) and into the WHO International Clinical Trials Registry Platform (ICTRP; www.who.int/ictrp/network/primary/en/) under shared catalogue number NTR6831. All participants provided written informed consent to participate in the study and to have their information obtained from treating physicians. |
| SARDINIA | SardiNIA | We thank all the volunteers who generously participated in this study and made this research possible.  This research was supported by the Intramural Research Program of the NIH, National Institute on Aging, with contracts N01-AG-1-2109 and HHSN271201100005C; by PB05 InterOmics MIUR Flagship Project and by grant FaReBio2011 “Farmaci e Reti Biotecnologiche di Qualità". |
| SASBAC cases / controls |  | This work was supported by grants from NIH (RO1-CA58427) and the Agency for Science, Technology and Research (A *STAR; Singapore). |
| SHIP | Study of Health in Pomerania | SHIP is part of the Community Medicine Research net of the University of Greifswald, Germany, which is funded by the Federal Ministry of Education and Research (grants no. 01ZZ9603, 01ZZ0103, and 01ZZ0403), the Ministry of Cultural Affairs as well as the Social Ministry of the Federal State of Mecklenburg-West Pomerania. Genome-wide data have been supported by the Federal Ministry of Education and Research (grant no. 03ZIK012) and a joint grant from Siemens Healthcare, Erlangen, Germany and the Federal State of Mecklenburg- West Pomerania. The University of Greifswald is a member of the Caché Campus program of the InterSystems GmbH. |
| SHIP-Trend | Study of Health in Pomerania - Trend | SHIP is part of the Community Medicine Research net of the University of Greifswald, Germany, which is funded by the Federal Ministry of Education and Research (grants no. 01ZZ9603, 01ZZ0103, and 01ZZ0403), the Ministry of Cultural Affairs as well as the Social Ministry of the Federal State of Mecklenburg-West Pomerania. The University of Greifswald is a member of the Caché Campus program of the InterSystems GmbH. |
| TWINGENE | TwinGene | TwinGene is part of the Swedish Twin Registry which is managed by Karolinska Institutet and receives funding through the Swedish Research Council under the grant no 2017-00641.  The Ministry for Higher Education; The Swedish Research Council (M-2005-1112); GenomEUtwin (EU/QLRT-2001-01254; QLG2-CT-2002-01254); NIH DK U01-066134; The Swedish Foundation for Strategic Research (SSF); Heart and Lung foundation no. 20070481 |
| TWINSUK | TwinsUK | TwinsUK is funded by the Wellcome Trust, Medical Research Council, European Union, Chronic Disease Research Foundation (CDRF), Zoe Global Ltd and the National Institute for Health Research (NIHR)-funded BioResource, Clinical Research Facility and Biomedical Research Centre based at Guy’s and St Thomas’ NHS Foundation Trust in partnership with King’s College London. |
| UK Biobank | UK Biobank | This research has been conducted using the UK Biobank resource under application numbers 871 and 9072 (Exeter) and 9797 (Cambridge). |
| VB | Val Borbera | We thank all the participants to the project, the San Raffaele Hospital MDs who contributed to clinical data collection, prof. Clara Camaschella who coordinated the data collection, Corrado Masciullo and Massimiliano Cocca for the database informatics.  The research was supported by funds from Compagnia di San Paolo, Torino, Italy; Fondazione Cariplo, Italy; Telethon Italy; Ministry of Health, Ricerca Finalizzata 2008 and 2011-2012 and Public Health Genomics Project 2010. |
| WGHS | Women's Genome Health Study | The WGHS is supported by the National Heart, Lung, and Blood Institute (HL043851 and HL080467) and the National Cancer Institute (CA047988 and UM1CA182913), with funding for genotyping provided by Amgen. NHLBI/NCI (Buring, Lee, PIs); Amgen (Chasman, Ridker, PIs). |
| WHI GARNET / WHIMS | Women's Health Initiative | The WHI program is funded by the National Heart, Lung, and Blood Institute, National Institutes of Health, U.S. Department of Health and Human Services through contracts HHSN268201600018C, HHSN268201600001C, HHSN268201600002C, HHSN268201600003C, and HHSN268201600004C. The authors thank the WHI investigators and staff for their dedication, and the study participants for making the program possible. A full listing of WHI investigators can be found at: https://s3-us-west-2.amazonaws.com/www-whi-org/wp-content/uploads/WHI-Investigator-Long-List.pdf |

**2) Studies included in Breast Cancer Association Consortium:**

| **Study acronym** | **Full study name** | **Acknowledgments and sources of funding** |
| --- | --- | --- |
| BCAC | Breast Cancer Association Consortium | We thank all the individuals who took part in these studies and all the researchers, clinicians, technicians and administrative staff who have enabled this work to be carried out. The COGS study would not have been possible without the contributions of the following: Kyriaki Michailidou, Andrew Lee, Craig Luccarini and the staff of the Centre for Genetic Epidemiology Laboratory, Javier Benitez, Anna Gonzalez-Neira and the staff of the CNIO genotyping unit, Jacques Simard and Daniel C. Tessier, Francois Bacot, Daniel Vincent, Sylvie LaBoissière and Frederic Robidoux and the staff of the McGill University and Génome Québec Innovation Centre, Sune F. Nielsen, Borge G. Nordestgaard, and the staff of the Copenhagen DNA laboratory, and Julie M. Cunningham, Sharon A. Windebank, Christopher A. Hilker, Jeffrey Meyer and the staff of Mayo Clinic Genotyping Core Facility.  BCAC is funded by Cancer Research UK [C1287/A16563, C1287/A10118], the European Union's Horizon 2020 Research and Innovation Programme (grant numbers 634935 and 633784 for BRIDGES and B-CAST respectively), and by the European Community´s Seventh Framework Programme under grant agreement number 223175 (grant number HEALTH-F2-2009-223175) (COGS). The EU Horizon 2020 Research and Innovation Programme funding source had no role in study design, data collection, data analysis, data interpretation or writing of the report. The breast cancer genome-wide association analyses were supported by the Government of Canada through Genome Canada and the Canadian Institutes of Health Research, the ‘Ministère de l’Économie, de la Science et de l’Innovation du Québec’ through Genome Québec and grant PSR-SIIRI-701, The National Institutes of Health (U19 CA148065, X01HG007492), Cancer Research UK (C1287/A10118, C1287/A16563, C1287/A10710) and The European Union (HEALTH-F2-2009-223175 and H2020 633784 and 634935). All studies and funders are listed in Michailidou et al (2017). |
| ABCFS | Australian Breast Cancer Family Study | ABCFS thank Maggie Angelakos, Judi Maskiell, Gillian Dite.  The Australian Breast Cancer Family Study (ABCFS) was supported by grant UM1 CA164920 from the National Cancer Institute (USA). The content of this manuscript does not necessarily reflect the views or policies of the National Cancer Institute or any of the collaborating centers in the Breast Cancer Family Registry (BCFR), nor does mention of trade names, commercial products, or organizations imply endorsement by the USA Government or the BCFR. The ABCFS was also supported by the National Health and Medical Research Council of Australia, the New South Wales Cancer Council, the Victorian Health Promotion Foundation (Australia) and the Victorian Breast Cancer Research Consortium. J.L.H. is a National Health and Medical Research Council (NHMRC) Senior Principal Research Fellow. M.C.S. is a NHMRC Senior Research Fellow. |
| ABCS |  | ABCS thanks the Blood bank Sanquin, The Netherlands.  The ABCS study was supported by the Dutch Cancer Society [grants NKI 2007-3839; 2009 4363]. |
| BBCC |  | The work of the BBCC was partly funded by ELAN-Fond of the University Hospital of Erlangen. |
| BCINIS |  | The BCINIS study would not have been possible without the contributions of Dr. K. Landsman, Dr. N. Gronich, Dr. A. Flugelman, Dr. W. Saliba, Dr. E. Liani, Dr. I. Cohen, Dr. S. Kalet, Dr. V. Friedman, Dr. O. Barnet of the NICCC in Haifa, and all the contributing family medicine, surgery, pathology and oncology teams in all medical institutes in Northern Israel. BIGGS thanks Niall McInerney, Gabrielle Colleran, Andrew Rowan, Angela Jones. |
| BREOGAN | BREast Oncology GAlician Network | The BREOGAN study would not have been possible without the contributions of the following: Manuela Gago-Dominguez, Jose Esteban Castelao, Angel Carracedo, Victor Muñoz Garzón, Alejandro Novo Domínguez, Maria Elena Martinez, Sara Miranda Ponte, Carmen Redondo Marey, Maite Peña Fernández, Manuel Enguix Castelo, Maria Torres, Manuel Calaza (BREOGAN), José Antúnez, Máximo Fraga and the staff of the Department of Pathology and Biobank of the University Hospital Complex of Santiago-CHUS, Instituto de Investigación Sanitaria de Santiago, IDIS, Xerencia de Xestion Integrada de Santiago-SERGAS; Joaquín González-Carreró and the staff of the Department of Pathology and Biobank of University Hospital Complex of Vigo, Instituto de Investigacion Biomedica Galicia Sur, SERGAS, Vigo, Spain.  The BREast Oncology GAlician Network (BREOGAN) is funded by Acción Estratégica de Salud del Instituto de Salud Carlos III FIS PI12/02125/Cofinanciado FEDER; Acción Estratégica de Salud del Instituto de Salud Carlos III FIS Intrasalud (PI13/01136); Programa Grupos Emergentes, Cancer Genetics Unit, Instituto de Investigacion Biomedica Galicia Sur. Xerencia de Xestion Integrada de Vigo-SERGAS, Instituto de Salud Carlos III, Spain; Grant 10CSA012E, Consellería de Industria Programa Sectorial de Investigación Aplicada, PEME I + D e I + D Suma del Plan Gallego de Investigación, Desarrollo e Innovación Tecnológica de la Consellería de Industria de la Xunta de Galicia, Spain; Grant EC11-192. Fomento de la Investigación Clínica Independiente, Ministerio de Sanidad, Servicios Sociales e Igualdad, Spain; and Grant FEDER-Innterconecta. Ministerio de Economia y Competitividad, Xunta de Galicia, Spain. |
| CBCS | Canadian Breast Cancer Study | CBCS thanks study participants, co-investigators, collaborators and staff of the Canadian Breast Cancer Study, and project coordinators Agnes Lai and Celine Morissette.  CBCS is funded by the Canadian Cancer Society (grant # 313404) and the Canadian Institutes of Health Research. |
| CCGP |  | CCGP thanks Styliani Apostolaki, Anna Margiolaki, Georgios Nintos, Maria Perraki, Georgia Saloustrou, Georgia Sevastaki, Konstantinos Pompodakis.  CCGP is supported by funding from the University of Crete. |
| CECILE |  | The CECILE study was supported by Fondation de France, Institut National du Cancer (INCa), Ligue Nationale contre le Cancer, Agence Nationale de Sécurité Sanitaire, de l'Alimentation, de l'Environnement et du Travail (ANSES), Agence Nationale de la Recherche (ANR). |
| CGPS |  | CGPS thanks staff and participants of the Copenhagen General Population Study. For the excellent technical assistance: Dorthe Uldall Andersen, Maria Birna Arnadottir, Anne Bank, Dorthe Kjeldgård Hansen. The Danish Cancer Biobank is acknowledged for providing infrastructure for the collection of blood samples for the cases.  The CGPS was supported by the Chief Physician Johan Boserup and Lise Boserup Fund, the Danish Medical Research Council, and Herlev and Gentofte Hospital. |
| CPSII |  | Investigators from the CPS-II cohort thank the participants and Study Management Group for their invaluable contributions to this research. They also acknowledge the contribution to this study from central cancer registries supported through the Centers for Disease Control and Prevention National Program of Cancer Registries, as well as cancer registries supported by the National Cancer Institute Surveillance Epidemiology and End Results program.  The American Cancer Society funds the creation, maintenance, and updating of the CPS-II cohort. |
| DIETCOMPLYF |  | The DietCompLyf study was funded by the charity Against Breast Cancer (Registered Charity Number 1121258) and the NCRN.  The University of Westminster curates the DietCompLyf database funded by Against Breast Cancer Registered Charity No. 1121258 and the NCRN. |
| ICICLE |  | ICICLE thanks Kelly Kohut, Michele Caneppele, Maria Troy.  ICICLE was supported by Breast Cancer Now, CRUK and Biomedical Research Centre at Guy’s and St Thomas’ NHS Foundation Trust and King’s College London. |
| KARBAC |  | Financial support for KARBAC was provided through the regional agreement on medical training and clinical research (ALF) between Stockholm County Council and Karolinska Institutet, the Swedish Cancer Society, The Gustav V Jubilee foundation and Bert von Kantzows foundation. |
| KARMA |  | KARMA and SASBAC thank the Swedish Medical Research Counsel.  The KARMA study was supported by Märit and Hans Rausings Initiative Against Breast Cancer. |
| KBCP |  | KBCP thanks Eija Myöhänen, Helena Kemiläinen.  The KBCP was financially supported by the special Government Funding (EVO) of Kuopio University Hospital grants, Cancer Fund of North Savo, the Finnish Cancer Organizations, and by the strategic funding of the University of Eastern Finland. |
| KCONFAB/ AOCS |  | kConFab/AOCS wish to thank Heather Thorne, Eveline Niedermayr, all the kConFab research nurses and staff, the heads and staff of the Family Cancer Clinics, and the Clinical Follow Up Study (which has received funding from the NHMRC, the National Breast Cancer Foundation, Cancer Australia, and the National Institute of Health (USA)) for their contributions to this resource, and the many families who contribute to kConFab.  kConFab is supported by a grant from the National Breast Cancer Foundation, and previously by the National Health and Medical Research Council (NHMRC), the Queensland Cancer Fund, the Cancer Councils of New South Wales, Victoria, Tasmania and South Australia, and the Cancer Foundation of Western Australia. Financial support for the AOCS was provided by the United States Army Medical Research and Materiel Command [DAMD17-01-1-0729], Cancer Council Victoria, Queensland Cancer Fund, Cancer Council New South Wales, Cancer Council South Australia, The Cancer Foundation of Western Australia, Cancer Council Tasmania and the National Health and Medical Research Council of Australia (NHMRC; 400413, 400281, 199600). G.C.T. and P.W. are supported by the NHMRC. RB was a Cancer Institute NSW Clinical Research Fellow. |
| MARIE |  | MARIE thanks Petra Seibold, Dieter Flesch-Janys, Judith Heinz, Nadia Obi, Alina Vrieling, Sabine Behrens, Ursula Eilber, Muhabbet Celik, Til Olchers and Stefan Nickels.  The MARIE study was supported by the Deutsche Krebshilfe e.V. [70-2892-BR I, 106332, 108253, 108419, 110826, 110828], the Hamburg Cancer Society, the German Cancer Research Center (DKFZ) and the Federal Ministry of Education and Research (BMBF) Germany [01KH0402]. |
| MCBCS |  | The MCBCS was supported by the NIH grants CA192393, CA116167, CA176785 an NIH Specialized Program of Research Excellence (SPORE) in Breast Cancer [CA116201], and the Breast Cancer Research Foundation and a generous gift from the David F. and Margaret T. Grohne Family Foundation. |
| MCCS | Melbourne Collaborative Cohort Study | The MCCS was made possible by the contribution of many people, including the original investigators, the teams that recruited the participants and continue working on follow-up, and the many thousands of Melbourne residents who continue to participate in the study.  The Melbourne Collaborative Cohort Study (MCCS) cohort recruitment was funded by VicHealth and Cancer Council Victoria. The MCCS was further augmented by Australian National Health and Medical Research Council grants 209057, 396414 and 1074383 and by infrastructure provided by Cancer Council Victoria. Cases and their vital status were ascertained through the Victorian Cancer Registry and the Australian Institute of Health and Welfare, including the National Death Index and the Australian Cancer Database. |
| MEC |  | The MEC was supported by NIH grants CA63464, CA54281, CA098758, CA132839 and CA164973. |
| MISS |  | The MISS study is supported by funding from ERC-2011-294576 Advanced grant, Swedish Cancer Society, Swedish Research Council, Local hospital funds, Berta Kamprad Foundation, Gunnar Nilsson. |
| MMHS |  | We thank the coordinators, the research staff and especially the MMHS participants for their continued collaboration on research studies in breast cancer.  The MMHS study was supported by NIH grants CA97396, CA128931, CA116201, CA140286 and CA177150. MSKCC is supported by grants from the Breast Cancer Research Foundation and Robert and Kate Niehaus Clinical Cancer Genetics Initiative. |
| NC-BCFR & OFBCR | Northern California Breast Cancer Family Registry (NC-BCFR) and Ontario Familial Breast Cancer Registry (OFBCR) | The Northern California Breast Cancer Family Registry (NC-BCFR) and Ontario Familial Breast Cancer Registry (OFBCR) were supported by grants U01CA164920 and U01CA167551 from the USA National Cancer Institute of the National Institutes of Health. The content of this manuscript does not necessarily reflect the views or policies of the National Cancer Institute or any of the collaborating centers in the Breast Cancer Family Registry (BCFR) or the Colon Cancer Family Registry (CCFR), nor does mention of trade names, commercial products, or organizations imply endorsement by the USA Government or the BCFR or CCFR. |
| NCBCS | Carolina Breast Cancer Study (NCBCS) | The Carolina Breast Cancer Study (NCBCS) was funded by Komen Foundation, the National Cancer Institute (P50 CA058223, U54 CA156733, U01 CA179715), and the North Carolina University Cancer Research Fund. |
| OFBCR |  | The OFBCR thanks Teresa Selander, Nayana Weerasooriya and Steve Gallinger. ORIGO thanks E. Krol-Warmerdam, and J. Blom for patient accrual, administering questionnaires, and managing clinical information. |
| PBCS |  | PBCS thanks Louise Brinton, Mark Sherman, Neonila Szeszenia-Dabrowska, Beata Peplonska, Witold Zatonski, Pei Chao, Michael Stagner.  The PBCS was funded by Intramural Research Funds of the National Cancer Institute, Department of Health and Human Services, USA. |
| PKARMA |  |  |
| PLCO |  | Genotyping for PLCO was supported by the Intramural Research Program of the National Institutes of Health, NCI, Division of Cancer Epidemiology and Genetics. The PLCO is supported by the Intramural Research Program of the Division of Cancer Epidemiology and Genetics and supported by contracts from the Division of Cancer Prevention, National Cancer Institute, National Institutes of Health. |
| SASBAC |  | The SASBAC study was supported by funding from the Agency for Science, Technology and Research of Singapore (A*STAR), the US National Institute of Health (NIH) and the Susan G. Komen Breast Cancer Foundation. |
| SBCS |  | SBCS thanks Sue Higham, Helen Cramp, Dan Connley, Ian Brock, Sabapathy Balasubramanian and Malcolm W.R. Reed.  The SBCS was supported by Sheffield Experimental Cancer Medicine Centre and Breast Cancer Now Tissue Bank. |
| SEARCH |  | We thank the SEARCH and EPIC teams.  SEARCH is funded by Cancer Research UK [C490/A10124, C490/A16561] and supported by the UK National Institute for Health Research Biomedical Research Centre at the University of Cambridge. The University of Cambridge has received salary support for PDPP from the NHS in the East of England through the Clinical Academic Reserve. |
| SISTER | The Sister Study | The Sister Study (SISTER) is supported by the Intramural Research Program of the NIH, National Institute of Environmental Health Sciences (Z01-ES044005 and Z01-ES049033). |
| SMC |  | The SMC is funded by the Swedish Cancer Foundation and the Swedish Research Council (VR 2017-00644) grant for the Swedish Infrastructure for Medical Population-based Life-course Environmental Research (SIMPLER). |
| UKBGS |  | UKBGS thanks Breast Cancer Now and the Institute of Cancer Research for support and funding of the Breakthrough Generations Study, and the study participants, study staff, and the doctors, nurses and other health care providers and health information sources who have contributed to the study. We acknowledge NHS funding to the Royal Marsden/ICR NIHR Biomedical Research Centre.  The UKBGS is funded by Breast Cancer Now and the Institute of Cancer Research (ICR), London. ICR acknowledges NHS funding to the NIHR Biomedical Research Centre. |
| USRT |  | The USRT Study was funded by Intramural Research Funds of the National Cancer Institute, Department of Health and Human Services, USA. |

**3) Further acknowledgments and sources of funding:**

Exeter:

Robin N. Beaumont is funded by the Wellcome Trust and Royal Society grant: 104150/Z/14/Z. Anna Murray is supported by the Wellcome Trust Institutional Strategic Support Award (WT097835MF). Katherine S. Ruth was supported by funding from the Gillings Family Foundation and MRCP2D. Jessica Tyrrell is supported by an Academy of Medical Sciences (AMS) Springboard award, which is supported by the AMS, the Wellcome Trust, GCRF, the Government Department of Business, Energy and Industrial strategy, the British Heart Foundation and Diabetes UK [SBF004\1079]. Andrew R. Wood is supported by the European Research Council grants: SZ-245 50371-GLUCOSEGENES-FP7-IDEAS-ERC and 323195. The authors acknowledge the use of the University of Exeter High-Performance Computing facility in carrying out this work.

*Mouse models of environmentally-induced low ovarian reserve:*

Catherine Aitken and Susan Ozanne are funded jointly by grants from the Academy of Medical Sciences, the Addenbrooke’s Charitable Trust, an Isaac Newton Trust/Wellcome Trust ISSF/ University of Cambridge Joint Research Grant and the MRC (MRC_MC_UU_12012/4). Susan Ozanne is a Member of the University of Cambridge MRC Metabolic Disease Unit.

*Mouse models of DNA damage response and ovarian ageing:*

Jazib Hussain is the recipient of an international PhD fellowship from the Punjab Educational Endowment Fund. Members of the Eva R Hoffmann were funded by an ERC Consolidator Grant (724718-ReCAP) and a Novo Nordisk Foundation Young Investigator Award (NNF15OC0016662). They acknowledge the support with histology from Pernille Sjølin Froh and Thi Cam Ha Nguyen at their Department. Andres Lopez-Contreras was funded by an ERC Starter grant (ERC-2015-STG-679068). Eva R Hoffmann and Andres Lopez-Contreras acknowledge core funding from the Danish National Research Foundation Center Grant (6110-00344B). Jeremy A. Daniels was supported by the Novo Nordisk Foundation (grant agreement NFF14CC0001). Satoshi H. Namekawa was funded by the National Institutes of Health (R01 GM098605). Claus Yding Andersen, Marie Louise Grøndahl, Kristina W. Olsen acknowledge core funding from Ferring and ReproUnion (www.reprounion.eu). Kristina W. Olsen was a recipient of a fellowship from Helsefonden (20-B-0334). Y Huang is a recipient of a fellowship of the China Scholarship Council (201607040048). Ignasi Roig was supported by the Spanish Ministerio de Ciencia e Innovación (BFU2013-43965-P, BFU2016-80370-P and PID2019-107082RB-I00).

**Individual study disclosures**

Joanne M Murabito discloses a consulting relationship with Merck as a guest lecturer.

All other authors and studies declared no conflict of interest.
